# A HYBRID INTEGRO-DIFFERENTIAL EQUATION AND NETWORK BASED MODEL OF EPIDEMICS

**DOI:** 10.1101/2024.04.04.24305342

**Authors:** Ted Duclos, Tom Reichert

**Affiliations:** Advisors, St. Augustine, Florida; Entropy Research Institute, Portland, Oregon

**Keywords:** Kermack and McKendrick integro-differential equations, Closed form solution, Epidemic management, Identification of outbreaks, Network Concepts

## Abstract

A closed form solution of the full Kermack and McKendrick integro-differential equations (Kermack and McKendrick 1927), called the KMES, is presented and verified. The solution is derived by combining network concepts with the integro-differential equations.

This solution has two parameters: one describing disease transmissibility and a second characterizing population interactions. The verified solution leads directly to useful, previously unknown, analytical expressions which characterize an epidemic. These include novel expressions for the effective reproduction number, time to peak in new infections, and the final size.

Using COVID -19 data from six countries, the transmissibility parameter is estimated and subsequently used to estimate the normalized contagiousness of an individual, a close approximation to viral shedding measured in infected persons. The population interaction parameter is estimated using the Google Residential Mobility Measure. With these parameter estimations, the KMES accurately projects case data from the COVID-19 pandemic in six countries over a 60-day period with R^2^ values above 0.85.

As to performance over longer periods, the KMES projects the Covid-19 total case data from the United States 21 days in advance over an 18-month period with a Mean Absolute Percentage Error of 4.1%. The KMES also accurately identifies the beginnings and peaks of outbreaks within multi month periods in case data from 4 countries.

## Introduction

Modern epidemiological modeling has its roots in the Kermack and McKendrick epidemic model, first published in 1927 (Kermack and McKendrick 1927). Since its publication, well over 10,000 authors have referenced and used this paper as a foundational starting point. However, as Diekmann points out in his insightful essay (2022 pg. 8), “…an incessant community-enforced misconception is that the paper is just about the very special case, the S-I-R (Susceptible, Infected, Recovered) model.” In the essay, he also elaborates on his suspicion that, despite the many references, the 1927 paper is rarely read by the citing authors. His concluding remarks decry the situation and request that the 1927 paper, a “…true gem…in which tremendous wisdom lies hidden.” (Diekmann 2022 pg. 9) be thoroughly read and its depths plumbed for further insights.

A closed form solution to Kermack and McKendrick’s integro-differential equations would very likely unlock additional wisdom, but the widespread use of SIR approximations (Breda et al 2021, Diekmann 2022 and Brauer 2008) and the recent publication of a discrete time model of the integro-differential equations (Diekmann 2021), are mute evidence that such a solution remains elusive. Lacking a solution, researchers have made prodigious efforts to extract wisdom from SIR approximations and produced what have become well-known formulas for the final size of epidemics, the notion of “flatten the curve” (DiLauro et al 2021), and various other refinements (e.g., Brauer 2008, Breda et al 2021, Hethcote 2000).

Unfortunately, approximations often incorporate unrealistic and unavoidable assumptions. For example, SIR approximations inherently assume that the affected population is well-mixed; an assumption that has been labelled as “unrealistically simple” (Brauer 2008, pg. 27). Therefore, lacking analytical alternatives, advances in the use of the approximations have tended to focus on approaches to minimize the effect of the assumptions.

One advanced approach, network-based stochastic models (e.g., Newman 2002, Diekmann et al 1998, and Youssef 2011) assume that epidemic dynamics can be projected by summing the collective probabilities that individuals will be infected with only a limited number of contacts; thereby avoiding the well-mixed assumption. These have been widely investigated and have produced estimations of outbreak threshold conditions and the final size of epidemics. However, while these are laudatory steps towards realistic characterizations of interactions, they do not yield the analytical expressions which make closed-form solutions so appealing and useful.

Inspired by Diekmann’s comments, we closely studied Kermack and McKendrick’s 1927 paper; and, by applying network model concepts, found a pathway to a closed-form solution. This approach frees the analysis from the assumptions inherent in the SIR approximations and produces analytical expressions with demonstrably practical application to the management of epidemics.

In this manuscript, we take up Diekmann’s call to action in three sections:

1. In Section 1 we present an analytical solution to the Kermack and McKendrick integro-differential equations and prove that it solves the equations. We then derive and explicate several analytical expressions characterizing epidemic dynamics. We call the solution the KMES (**<u>K</u>**ermack and **<u>M</u>**cKendrick **<u>E</u>**quation **<u>S</u>**olution).
2. In Section 2, using data compiled from several countries during the COVID-19 pandemic, we demonstrate that a transmissibility and population interaction parameter can be deduced from early pandemic data, combined with the independently sourced Google Residential Measure (Google 2023). With these parameters, the KMES not only accurately projects the course of the COVID-19 pandemic in the six sampled countries, but also provides an estimate of the normalized contagiousness of an individual.
3. In Section 3 we derive additional expressions which can be used to determine the actions necessary to diagnose, control, and end an epidemic. We demonstrate the use of these expressions with the United States COVID-19 case data from April 2020 to November 2021; and accurately project the total case data 21 days in advance over 18 months. Lastly, we demonstrate the ability to detect in advance the major outbreaks that occurred during the pandemic in 4 sampled countries.

### Section 1: A Solution to the Kermack and McKendrick Equations

(Note: We use the following equation notation, (X, SY-Z), where X is the equation number in the body; and if the equation is used in a supplement, SY is the supplement number and Z is the number of the equation in the supplement). A list of all equations is provided in Supplement 4.

#### Definition of Terms

We model the population affected by the epidemic as a network with *N*_*P*_ vertices. The contacts of each person/vertex in *N*_*P*_ which can transmit an infection are the edges. Within this network, we define *N*_*P*_ as the total number of people who can possibly become infected during the epidemic; *S*(*t*), as the subpopulation that has not yet become infected; *N*(*t*) as the subpopulation that is currently or has previously been infected; *I*(*t*) as the infectious individuals within *N*(*t*); and R(*t*) as the total recovery of individuals within *N*(*t*). *S*(0) is the number of uninfected people in *N*_*P*_ at the epidemic start, *S*(∞) is the number of uninfected people at the end of the epidemic, and *I*(0) is the number of infectious individuals at the epidemic start. Therefore, *N*_*P*_ = *S*(*t*) + *N*(*t*), *N*(*t*) = *I*(*t*) + *R*(*t*), and *N*(0) = *I*(0). We further assume that for our modelling purposes, *N*_*P*_ is a constant; and that once infected and recovered, people cannot become reinfected.

Since not everyone who is infected can transmit the disease, we distinguish three subgroups of infected people as follows:

1. People who have just been infected but are not yet shedding virus are latent infectious.
2. People who are shedding the virus and are in contact with susceptible people are infectious.
3. People who may be shedding the virus but have no contact with susceptible people are part of the recovered group *R*(*t*).

In our solution, persons in groups 1 and 2 are part of *I*(*t*), collectively known as infectious, and persons in groups 2 and 3 are considered contagious. Members of all three groups are part of the ever-infected population, *N*(*t*). To avoid any ambiguities in these definitions, we clarify them further using two thought experiments.

In the first thought experiment, imagine that a person has just infected a particular person. In this case, the infecting person cannot reinfect that newly infected person nor can the newly infected person infect their infector. In our model, such people are vertices in a network where the edges are contacts which could transmit an infection and we assume that if the frequency of contact or the number of contacts (edges) between people does change, these do so slowly when compared to the rate at which people become infected.

Additionally, we assume that infectors remain in durable potentially infectious contact with the people they have infected. Consequently, the infectiousness, but not the degree, of infectors <u>diminishes</u> with each infection they cause; and, in symmetry, the extent to which they are unable to infect others, their recovery increases. This change in infectiousness does not affect their level of contagiousness which is considered purely a function of the disease.

In the second thought experiment, as described in Diekmann (2022), during an infection, the quantity and quality of the infectious agent within any infected person, the so-called “viral load”, will rise and fall with time. Since the contagiousness of an infected person will also vary in synchrony, we postulate that the level of infectiousness will approximately track this variation in the contagiousness. Those persons whose infection has passed the maximum point of contagiousness will become increasingly less infectious; and, correspondingly more recovered. We say “approximately” when discussing this relationship, because infectiousness and contagiousness are not equivalent, as the notions developed in the definitions and first thought experiment make clear.

#### Kermack and McKendrick’s Model Structure

Kermack and McKendrick (1927) derived their integro-differential equations by imagining *N*(*t*) as the sum at time t of incremental subpopulations which had been infected at prior times *t* − *θ*; where *θ* specifies the time since infection of each subpopulation. An array illustrating the relationship between time and *θ* underpinning their equations is presented in Figure 1. As Figure 1 shows, since *θ* has the units of time, each increment of *θ* is designated as Δ*t*. Both t and *θ* must be ≥ 0; and because *θ* ≤ *t* and Δ*θ* = Δ*t*, the array is square.

**Figure 1.**
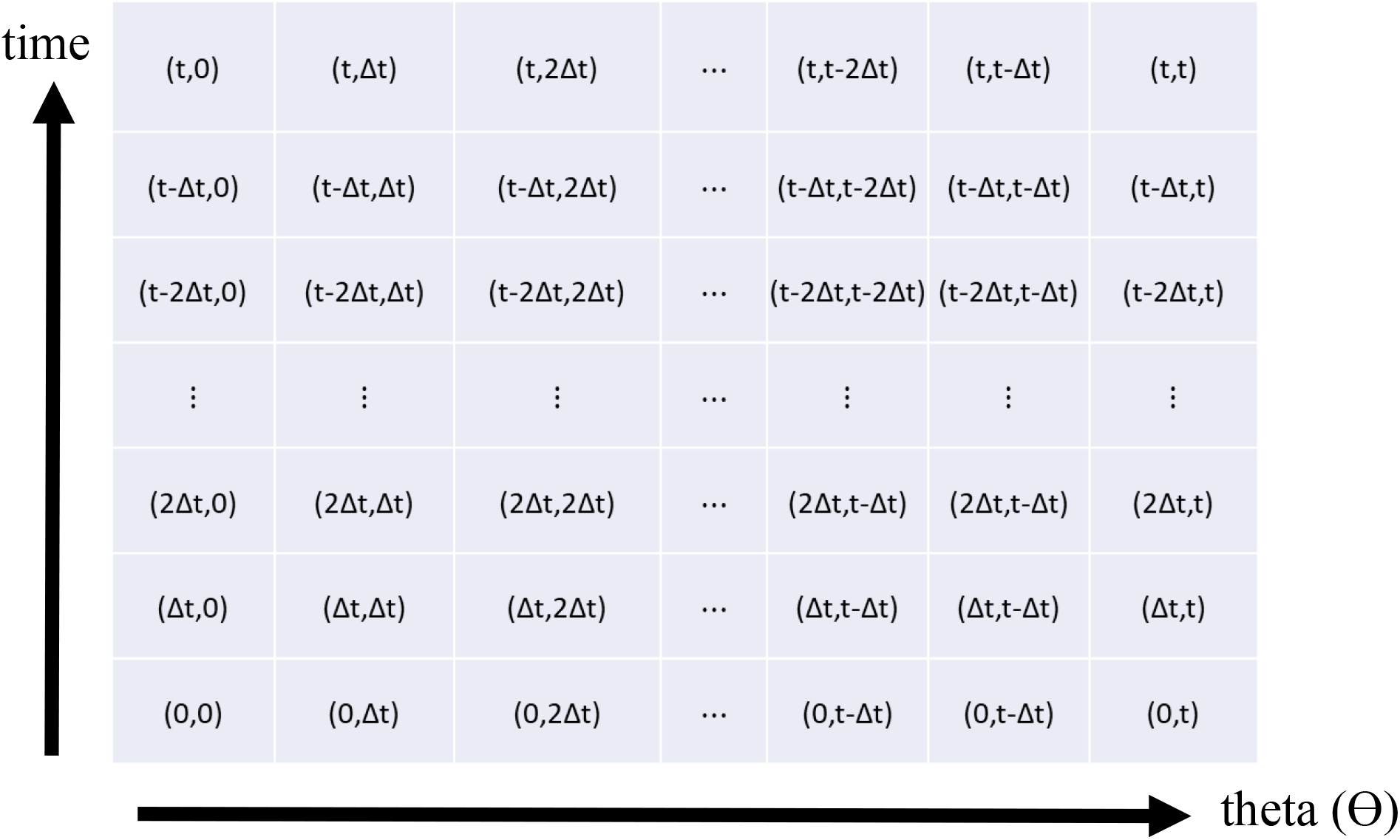
Array showing time and theta. A new row on top and a new column to the right is added with each Δ*t* increment of time.

Designating each incremental subpopulation infected at a time *t* − *θ* as a “*θ*-group”, Kermack and McKendrick’s analysis tracked the progress of each *θ*-group through both time and *θ*. Since Δ*θ* = Δ*t*, this means each group started in the left most column of the *t* − *θ* row in Figure 1 and moved diagonally upwards to the right thereafter. Their integro-differential equations were created by first summing each row over *θ* and then taking the limit of those summations as Δ*θ* → 0.

We adapt the format of Kermack and McKendrick’s equations to our previously defined population variables using a specific notation. Unless otherwise specified, all dependent variables and parameters are assumed to depend on both time, t, and *θ* (e.g., *I*(*t, θ*)). Additionally, if a population variable representing a portion of the population is designated as a function of time alone, then it is to be understood that that variable has been integrated over *θ* from 0 → *t*. For instance, 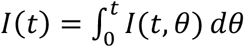. However, if we designate a parameter as solely a function of time or *θ*, then that parameter is to be considered constant over the alternate temporal dimension, (i.e, *θ* or time) respectively. The population variables used in this manuscript are S, I, N, and R; and the parameters are identified as they are introduced to the analysis.

With the conceptualizations developed above, the Kermack and McKendrick integro-differential equations in terms of time, *θ*, and population variables are:

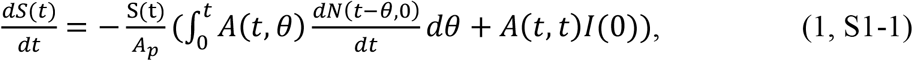

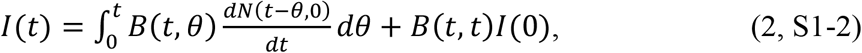

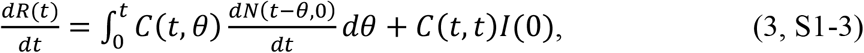

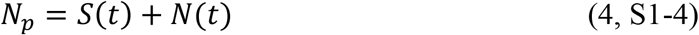

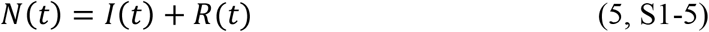

where 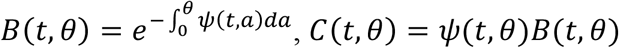, and *A*(*t, θ*) = *φ*(*t, θ*)*B*(*t, θ*). Kermack and McKendrick (1927, p. 703) defined *φ*(*t, θ*) as “the rate of infectivity at age *θ*”, and *ψ*(*t, θ*) as “the rate of removal” of the infected population to the recovered population and we continue these associations. *N*_*P*_ is the total population, *A*_*P*_ is the area that contains *N*_*P*_ and, as previously defined, *θ* is the time since infection of any member of the population *N*(*t*). We also note that in Equations 1, 2, and 3, 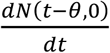 is the new infections at time *t* − *θ*.

#### The Solution

In Supplement 1, we derive a solution to Equations 1-5 and call it the KMES (**<u>K</u>**ermack and **<u>M</u>**cKendrick **<u>E</u>**quation **<u>S</u>**olution). The KMES is stated as,

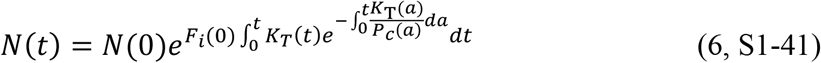

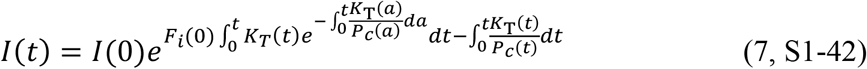

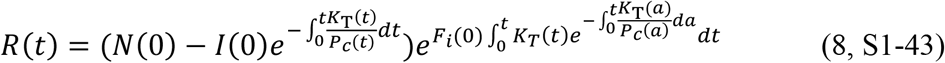

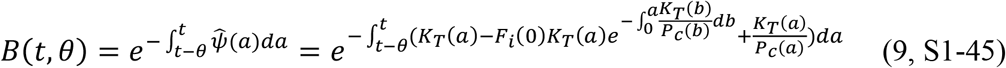

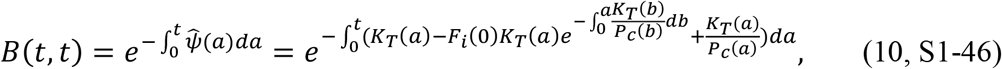

where the integration limits of the exponential within *B*(*t, θ*) in Equation 9 have been restated as the interval from *t* − *θ* to *t*, instead of the interval, 0 to *θ*, specified by Kermack and McKendrick. This interval, *t* − *θ* to t, with a length of *θ*, extends from the time of infection to the current time, and is the same time interval referenced in Kermack and McKendrick’s formula for *B*(*t, θ*). A detailed explanation of the basis for this crucial restatement is given in Supplement 1 (Equations S1-15 through S1-24). The parameters, 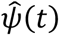, *K*_*T*_(*t*), *P*_*c*_(*t*), and *F*_*i*_(*t*), are described in the following paragraphs.

The derivation of the solution uses the weighted averages of *φ*(*t, θ*) and *ψ*(*t, θ*),

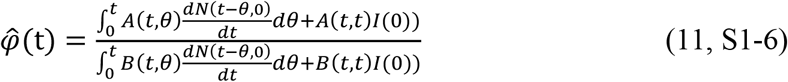

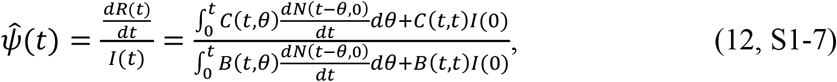

the formulation of which utilize the forms of *A*(*t, θ*) = *φ*(*t, θ*)*B*(*t, θ*), and *C*(*t, θ*) = *ψ*(*t, θ*)*B*(*t, θ*). It was also convenient to define the parameter, *K*_*T*_ (*t*), in terms of 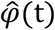 and 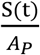 as,

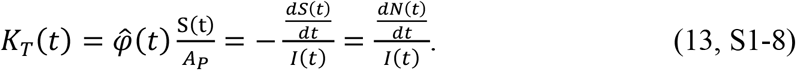

From its form, *K*_*T*_ (*t*) must be a transmission rate with the 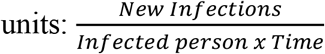, and is affirmed as a property of the disease in Section 2. Equation 13 embodies both the notion that all new infections are caused by contact with the currently infectious, and that 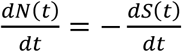 because all new infections must come from the susceptible population. Also, to address the matter of a latency period, this effect, if present in the disease, is manifested as a lowering of the value of both 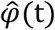 and *K*_*T*_(*t*) early in the epidemic, until there are enough contagious infections to offset the delay associated with the latency.

As previously stated, we assume the population can be represented as a network where the individuals are the vertices, and the edges are the contacts capable of transmitting the disease. In deriving the solution, we further assumed that, within this population, there is a subnetwork wherein only those contacts that already have, or eventually will, pass on an infection are the edges of this subnetwork. We also assume that the mean number of these edges per person, which we call *P*_*c*_(*t*), is the self-weighted mean degree of this subnetwork minus 1. We subtract 1 because a person cannot infect their infector.

As defined, *P*_*c*_(*t*) is equivalent to the basic reproduction number, *R*_0_, a concept used in conventional modelling terminology; but since we utilize it in an unconventional manner, we refer to it as *P*_*c*_(*t*) throughout the manuscript. To affirm this equivalence, wherever possible, we highlight where and in what way *P*_*c*_(*t*) exhibits characteristics conventionally thought of as manifest in *R*_0_.

The use of *P*_*c*_(*t*) allows the analysis to proceed by only tracking the number of infection-transmitting interactions that infected people have with susceptible people, and for this number to vary independently from the <u>total</u> number of susceptible people remaining within *N*_*P*_. This frees the model from the constraint of the “well-mixed” assumption wherein all the infected people are assumed to be in contact with all the susceptible people at any given time.

This also frees the analysis from having to track <u>all</u> interactions and the probabilities that they will transmit an infection between infected people and their contacts. Instead, the specification of *P*_*c*_(*t*) acknowledges that an epidemic progresses with the passing of infections between infectious and susceptible people, which occurs *P*_*c*_(*t*) times per infectious person, at a rate *K*_*T*_(*t*). We also hypothesize that *P*_*c*_(*t*) is related to the population behavior and demonstrate this in Section 2. The conceptualization of *P*_*c*_(*t*) is further expanded in Supplement 1.

The last important parameter in the solution, *F*_*i*_(*t*), the fraction of *N*(*t*) that is infected, is defined as,

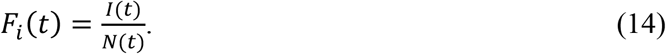

This parameter enables us to transform Equation 12 into a useful, alternative expression for 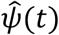 in terms of *K*_*T*_(*t*), *P*_*c*_(*t*), and *F*_*i*_(*t*),

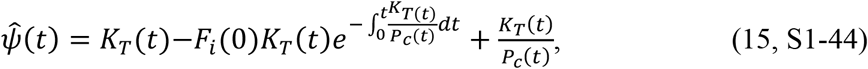

#### Proof of the Solution

We now substitute the solution into Equations 1 through 5 to prove it is indeed a solution. We start by substituting Equation 15 into Equation 7 to obtain,

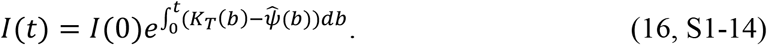

Also, by differentiating Equation 6,

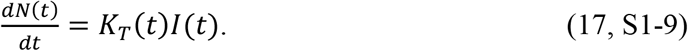

Since we know that 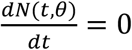 for all *θ* > 0, and therefore 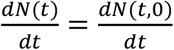, we can use this relationship in Equation 17, and substitute the result along with Equations 16, 9 and 10 into Equation 2. Keeping in mind that *dθ* = *dt*, we subsequently rewrite Equation 2 as,

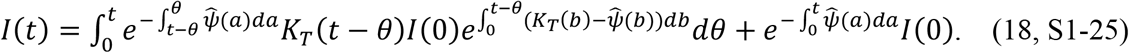

which can be simplified to,

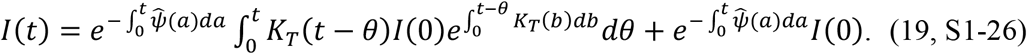

Since 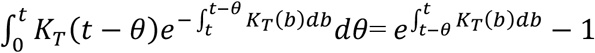, the middle integral in the first term in Equation 19 can be evaluated, and Equation 19 collapses to:

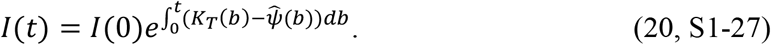

Equation 20 is identical to Equation 16, thereby proving that Equation 16 (and hence Equation 7 in the KMES) is a solution to Equation 2.

The proof of solution for Equation 1 is similar and begins by substituting Equations 6, 17, 12, 9 and 10 into Equation 1 to obtain,

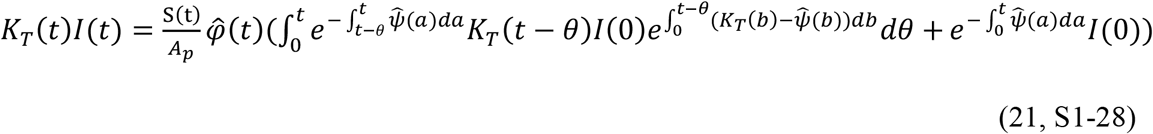

Using Equations 18 and 20, and noting that 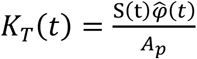, Equation 21 can be simplified to,

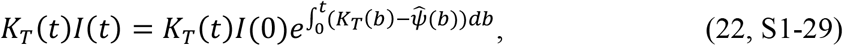

If we then substitute Equations 15 and 17 into 22, we find the following expression,

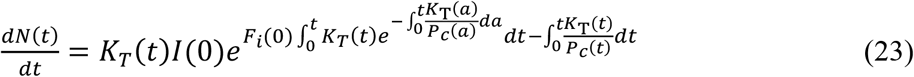

After dividing both sides of Equation 23 by Equation 6, and then integrating, we obtain,

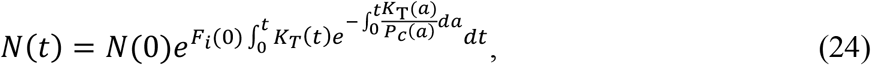

which is identical to Equation 6 proving that we have a solution to Equation 1.

Lastly, from Equation 12 we know that 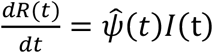, and since, when we differentiate Equation 8, we find the same expression, Equation 8 is a solution to Equation 3.

#### Simplified Expressions

The expressions for *N*(*t*), *I*(*t*), and *R*(*t*) can be rewritten in simplified, more intuitive forms. The first of these is what we call the Step Response form, found by using Equation 10 to rewrite *N*(*t*), *I*(*t*), and *R*(*t*) as,

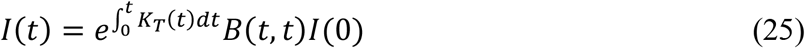

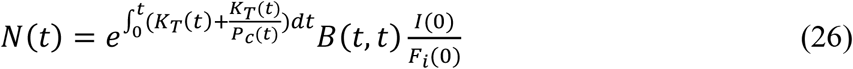

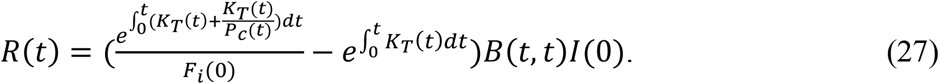

Since *B*(*t, t*) is the time varying infectiousness input to the original infected group, *I*(0), the exponential expressions, 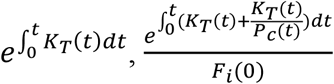, and 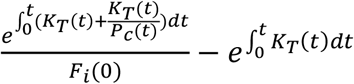, are the step response functions to this input.

As a sensibility check, Equation 25 shows that if there were no recovery, that is, if *B*(*t, t*) = 1, the infections would grow exponentially, at a rate determined by the transmissibility of the agent, until the entire population was infected, an intuitive and reasonable result. This also makes clear that, even with a finite recovery period, with large enough population interactions, *P*_*c*_(*t*), everyone in the initially affected population, *S*(0), could eventually become infected.

As a final simplification, if we can assume that both *K*_*T*_(*t*) and *P*_*c*_(*t*) in Equations 6 to 8 can be constant for a period, we arrive at the following expressions,

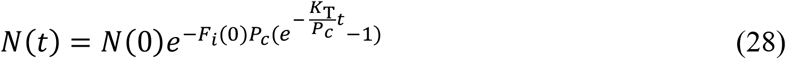

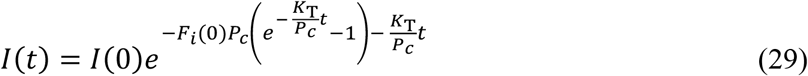

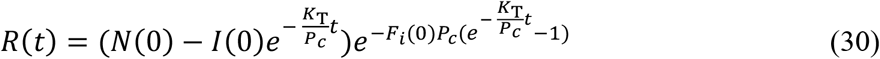

#### Expressions Describing an Epidemic

Beyond sensibility checks, the availability of a closed form solution opens a pathway to the derivation of expressions useful in epidemic description and management. We illustrate several of these here using the simplifying assumption that *K*_*T*_ and *P*_*c*_ are constants, which allows the nature of the expressions to be more easily seen and understood.

#### Effective and Basic Reproduction Number, R_eff_ and R_0_

Using Equation S1-40 from Supplement 1, we can write an expression for the average number of susceptible people remaining within *P*_*c*_ as,

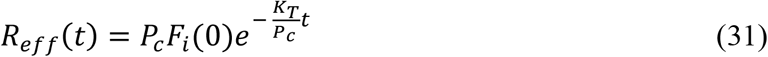

We call *R*_*eff*_(*t*) the effective replication number because it is the average number of contacts per infectious person that they will still infect as long as *P*_*c*_ remains constant.

In Supplement 1, while deriving the solution, we also develop Equation S1-10, an expression useful in diagnosing whether an epidemic is growing,

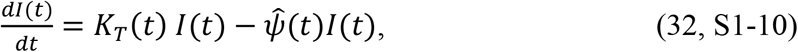

which, by using Equation 15 and 31, can be rewritten as,

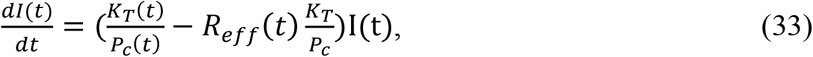

Therefore, if *R*_*eff*_(*t*) > 1, the epidemic is growing, and reciprocally, if *R*_*eff*_(*t*) < 1, the epidemic is declining. This means that *R*_*eff*_(*t*) is an indicator of the epidemic’s direction, and, as we show in Section 3, is a diagnostic metric for potential use in public health decision making. Lastly, we note that if *F*_*i*_(0) = 1, then *R*_*Eff*_(0) = *P*_*c*_, corroborating that *P*_*c*_ is equivalent to the Basic Reproduction Number, *R*_0_.

#### Time to Peak New Infections and Peak Size

Setting *R*_*Eff*_(*t*) = 1 in Equation 31 yields the time when *I*(*t*) begins to decline:

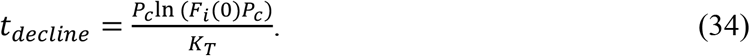

Likewise, by differentiating both sides of Equation 28, the expression for *N*(*t*), twice and equating the result to zero, we find the time when 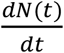 will be maximal,

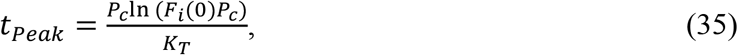

where *t*_*Peak*_ = the time to the peak of new infections. As it should, the peak in new cases coincides with the start of the decline of infections.

Equation 35 projects that when social interventions are stronger (smaller *P*_*c*_), the time to the peak will <u>always</u> be shorter, the opposite of the SIR models’ projection of this relationship. We highlight this qualitative difference between the projections of the KMES and SIR models because, as we show in Section 3, the data from the Covid-19 pandemic supports the KMES projection.

Equation 34 can be substituted into Equation 29 to find the peak value of the infections,

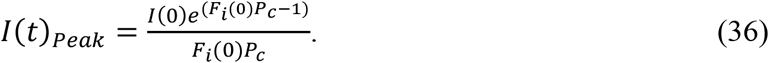

During an epidemic, Equation 36 can be used to estimate the maximum level of infections and derivative need for office visits or hospitalizations that may occur in the future.

#### Final Size

In the limit, as *t* → ∞, Equation 28 generates an expression for the final size,

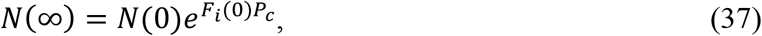

and the time it would take for *N*(∞) = *N*_*P*_ is,

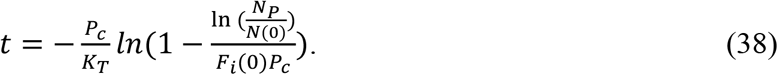

Therefore, as suggested by the Step Response form of the solution, if *P*_*c*_ is large enough for a long enough period, the KMES projects that it is possible, at some point in time, for *N*(*t*) to equal *N*_*P*_. The criteria for this to occur is,

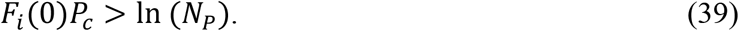

This result differs from Kermack and McKendrick’s conclusion that the entire susceptible population cannot be infected, a result that has achieved a position of some prominence within the epidemiological modelling community. Therefore, we feel compelled to defend our conclusion by pointing out that our path to this result was completely straightforward. To wit, by applying the definitions of *K*_*T*_(*t*) and 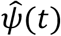, Equations 28 through 30 can be demonstrated to be solutions to the integro-differential equations 1 through 5. Equations 37 and 38 are then directly derived from Equation 28, leading to the inescapable conclusion that the integro-differential equations allow for the entire population to become infected.

Kermack and McKendrick did not derive a solution and therefore could not derive expressions equivalent to Equations 37 and 38. Rather, their conclusion rested upon the assumption that their function *A*, defined as 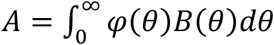, is always finite. This assumption, however, was unjustified because *φ*(*θ*) is the rate of new infections ***Per susceptible***; and as the number of susceptibles approaches zero, *φ*(*θ*), and consequently *A*, will tend towards infinity. Had this observation been made by Kermack and McKendrick, it would surely have led them to the same conclusion.

As an additional bridge to conventional notions of the final size, Equation 37 can be deduced from a special case of the SIR model final size equation. From Brauer (2008) this equation is,

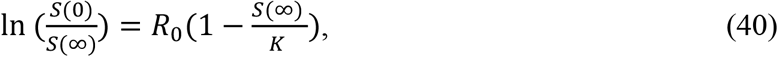

where *K* ≈ *S*(0).

Utilizing our prior definition of the relationships between *N*_*P*_, *S*(*t*), and *N*(*t*),

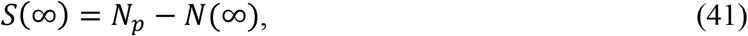

if we then substitute this into Equation 40 and use *K* ≈ *S*(0), we obtain:

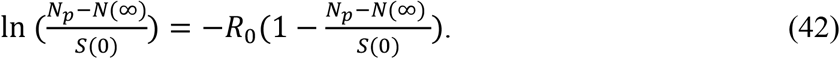

In a special case, if the final size *N*(∞) equals *N*_*P*_ − 1, which is equivalent to when *S*(∞) = 1, we can substitute this into Equation 42, exponentiate, and since 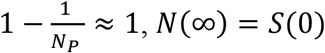, and *N*(0)=1, arrive at,

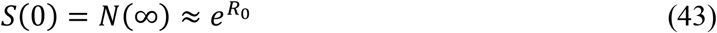

Using the equivalency *R*_0_ = *P*_*c*_, if *F*_*i*_(0) = 1, then Equation 43 is identical to Equation 37.

The equivalency range between Equation 40 and Equation 37 can be expanded beyond the special case by applying the following logic. First, Equation 40 is only valid for *S*(∞) > 0 because the SIR model equation it is derived from, 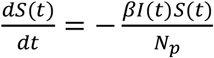, can only be solved for *S*(*t*) when *S*(*t*) > 0. Second, the magnitude of *S*(∞) depends on the value chosen for *S*(0). Therefore, when modeling an epidemic in a population, *N*_*A*_, a subpopulation, *N*_*P*_ = *S*(0) + *N*(0), relevant to the epidemic can be chosen such that *N*(∞) + 1 = *N*_*P*_. With this choice, Equation 43, and by extension, Equation 37 will be the same final size equation. For those situations where *F*_*i*_(0)*P*_*c*_ > ln (*N*_*A*_), then equation 40 is not valid and Equation 37 projects that the whole population will be infected. While this analysis is purely hypothetical, it demonstrates a conceptual bridge between the SIR approximations and the KMES solution.

### Section 2: Projections of Pandemic Data Using the KMES

In this section, we first demonstrate methods for estimating *K*_*T*_(*t*) and *P*_*c*_(*t*) using COVID-19 epidemic data, and then, armed with these estimates, we use the KMES to project the normalized contagiousness in individuals with COVID-19 and the progression of cases in six different countries.

#### Estimation of *K*_*T*_(*t*)

Other than where *K*_*T*_(*t*) arises in the model, we have no other a priori information about it. Therefore, we assume that *K*_*T*_(*t*) is a parameter associated with the disease and possibly constant for intervals in which the infectious agent does not change. We then begin the search for the value of *K*_*T*_(*t*) by stating some basic assumptions regarding the relationship between *P*_*c*_(*t*) and the population density.

Because the population of a country typically is only physically present within ∼1% of the land in a region (Ritchie and Roser 2019), we can define an “effective area” parameter, *A*_1_(*t*), where 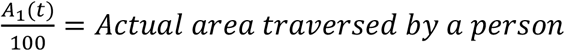. If we assume in the initial stages of the epidemic that *P*_*c*_(*t*) is a function of population density, and that an individual’s infectious mobility extends over an average effective area per unit of time, we can then write an expression in terms of this area:

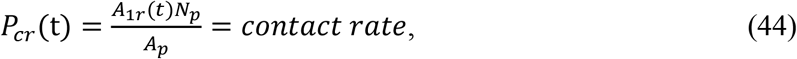

where *P*_*cr*_(*t*) is the contact rate associated with *P*_*c*_ (see Equation S1-32 in Supplement 1), *N*_*P*_ is the entire population of the region containing the infection, *A*_*P*_ is the area of the region, 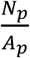 is the population density, and *A*_1*r*_(*t*) is the effective area per unit time that an individual infectiously inhabits.

We can then define *A*_1_(*t*) as,

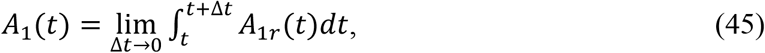

In specifying this area, we also assume, like the assumption that we made for *P*_*c*_(*t*) in Section 1, that changes in the value of *A*_1_(*t*) occur slowly in time. We can now write a new expression for 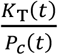,

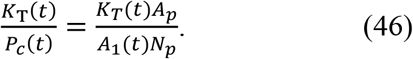

Assuming *N*(0) and *F*_*i*_(0) both are equal to 1, and that *K*_*T*_(*t*), *P*_*c*_(*t*), and *A*_1_(*t*) are constants in the initial stages of the epidemic, substituting Equation 46 into the equation for *N*(*t*) (Equation 28) produces an expression,

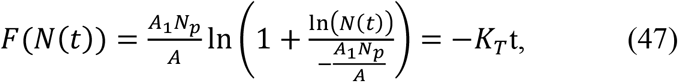

which predicts that if *K*_*T*_ and *A*_1_ are well approximated as constants, then *F*(*N*(*t*)) is a linear function of time. Excepting *K*_*T*_ and *A*_1_, all the quantities in Equation 47 can be found and are listed in Table 1 for six countries in the time before containment measures were enacted.

**Table 1.**
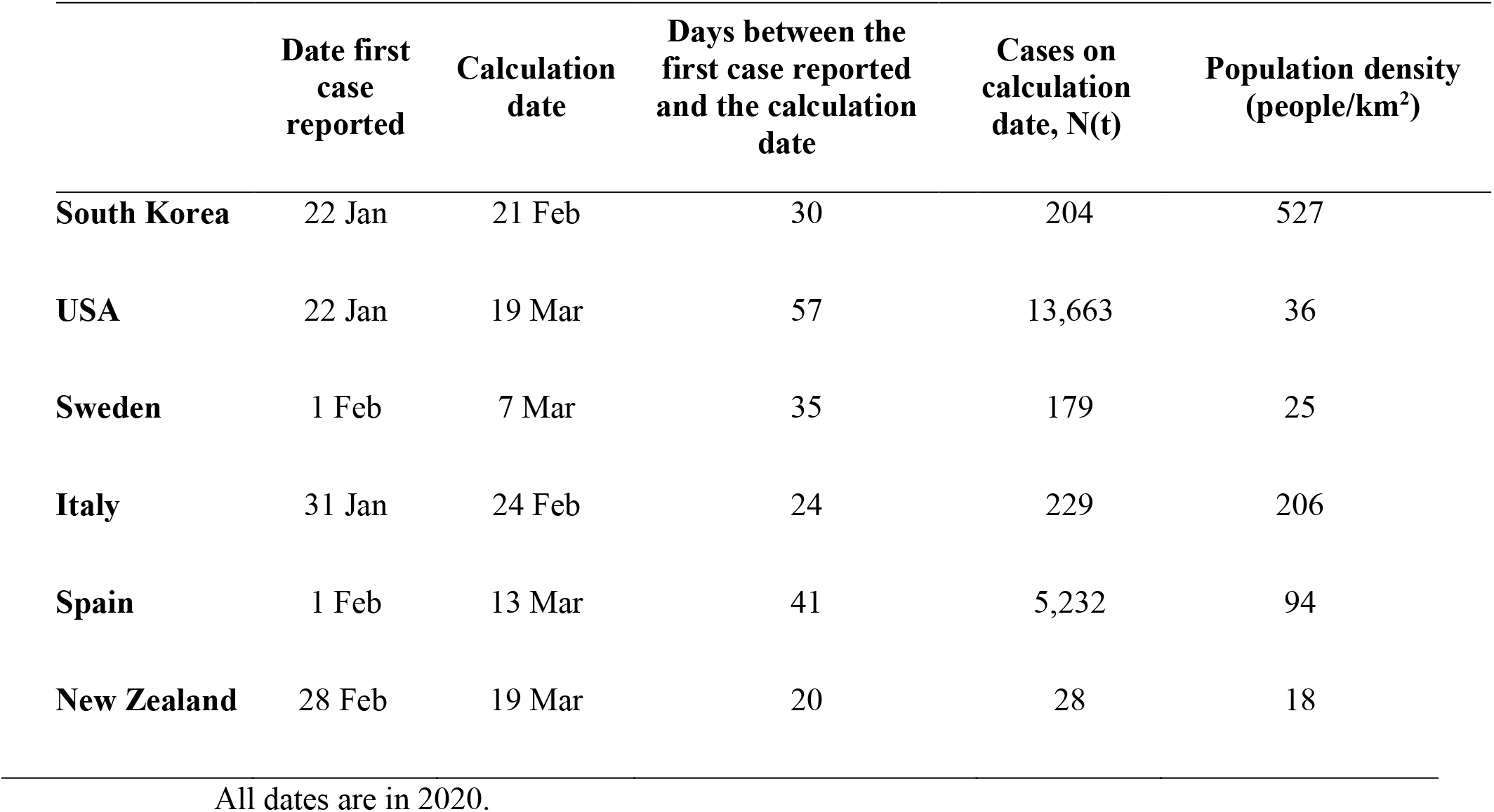
Initial COVID-19 pandemic data and population densities for various countries ((Roser et al 2021), case and date data; (Worldometers 2021, population density data)

|  | Date first case reported | Calculation date | Days between the first case reported and the calculation date | Cases on calculation date, N(t) | Population density (people/km<sup>2</sup>) |
| --- | --- | --- | --- | --- | --- |
| **South Korea** | 22 Jan | 21 Feb | 30 | 204 | 527 |
| **USA** | 22 Jan | 19 Mar | 57 | 13,663 | 36 |
| **Sweden** | 1 Feb | 7 Mar | 35 | 179 | 25 |
| **Italy** | 31 Jan | 24 Feb | 24 | 229 | 206 |
| **Spain** | 1 Feb | 13 Mar | 41 | 5,232 | 94 |
| **New Zealand** | 28 Feb | 19 Mar | 20 | 28 | 18 |

All dates are in 2020.

We then applied Equation 47 to six different countries using the data in Table 1 and through a process of iteration, determined that the best value for *A*_1_ to create the straight line plotted in Figure 2 was 0.48 km^2^. Since *A*_1_ was the effective area, the estimated actual area was 4800 m^2^, or a 70 m x 70 m square, a plausible area for a person to traverse in a day.

**Figure 2.**
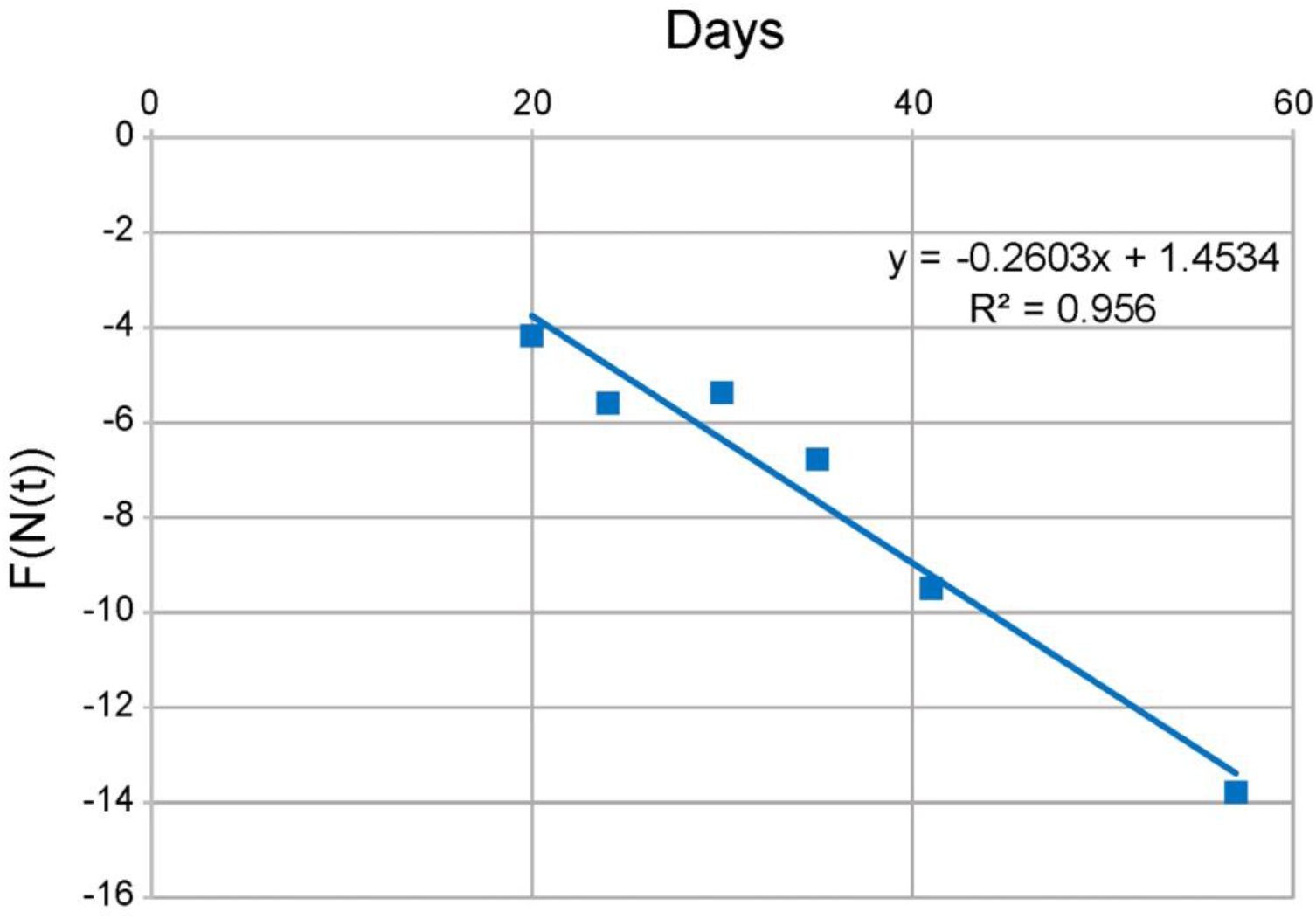
Verification that *K*_*T*_ may be the same for all countries. The data from Table 1 is plotted using Equation 47 and *A*_1_ = 0.48 *km*^2^. Each data point corresponds to a different country. The value of *K*_*T*_ is the negative of the slope of the line, and *K*_*T*_ is closely approximated everywhere by *K*_*T*_ ≈ 0.26

As noted in Figure 2, the line fit to the data points has an *R*^*2*^ = 0.956 and slope of 0.26 (with a 95% confidence interval, CI, from 0.4 to 0.13), the putative value of *K*_*T*_. This is strong support for the assumption that *K*_*T*_ and *A*_1_could be constant in the initial stages of the pandemic across all the sampled countries and thus *K*_*T*_ can be taken as a parameter of the disease.

#### Contagiousness

At the beginning of Section 1, in the thought experiments, we explained that contagiousness is a notion separate from infectiousness. Since contagiousness is also assumed to be a function of the disease, extracting an expression for contagiousness from the KMES should offer insight into the average dynamics of the disease progression in individuals. Furthermore, if the contagiousness of individuals can be measured, then this expression could serve as an independent benchmark against which these measurements could be compared.

The step response structure of the KMES suggests that such an expression should exist, hence *B*(*t, t*) is a reasonable starting point because it describes the evolution of infectiousness in the initially infected population. Beginning with Equation 10, in an imaginary scenario, if we assume that *K*_*T*_(*t*) is a constant; and that *P*_*c*_ = *I*(0) = 1 (i.e., there was only one initial infection, and this initially infected person contacted, on average, only one other person), the infectiousness of the initially infected person would be,

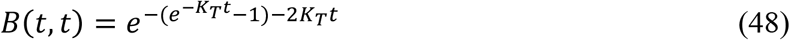

Equation 48 describes the capacity of the initially infected person to infect, a quantity which is dependent on the portion of contacts that remain infectable as well as the person’s contagiousness. Therefore, since the available number of infectable people in *P*_*c*_ is equal to *R*_*Eff*_, dividing *B*(*t, t*) by *R*_*Eff*_ (divide Equation 48 by Equation 31) extracts the contagiousness. This yields, for *P*_*c*_ = 1,

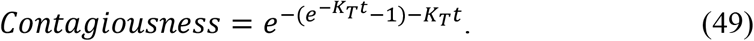

which is only dependent on *K*_*T*_, confirming the assumption that *K*_*T*_ is a parameter of the disease.

Figure 3, a plot of Equation 49, was created using the nominal and 95% CI values for *K*_*T*_ generated from Figure 2, and the shapes of the curves in the figure depict the time course of an individual’s estimated contagiousness. As can be seen in the figure, these shapes are highly similar to the expected time history of an individual’s viral load. Indeed, the nominal curve in the figure appears to project that an individual infected with COVID-19 would become contagious about 5 days before the peak; and have minimal contagiousness approximately 10 days after the peak. This timetable coincides closely with reported measured values describing viral shedding (Puhach et al 2023, Figures 2 and 3).

**Figure 3.**
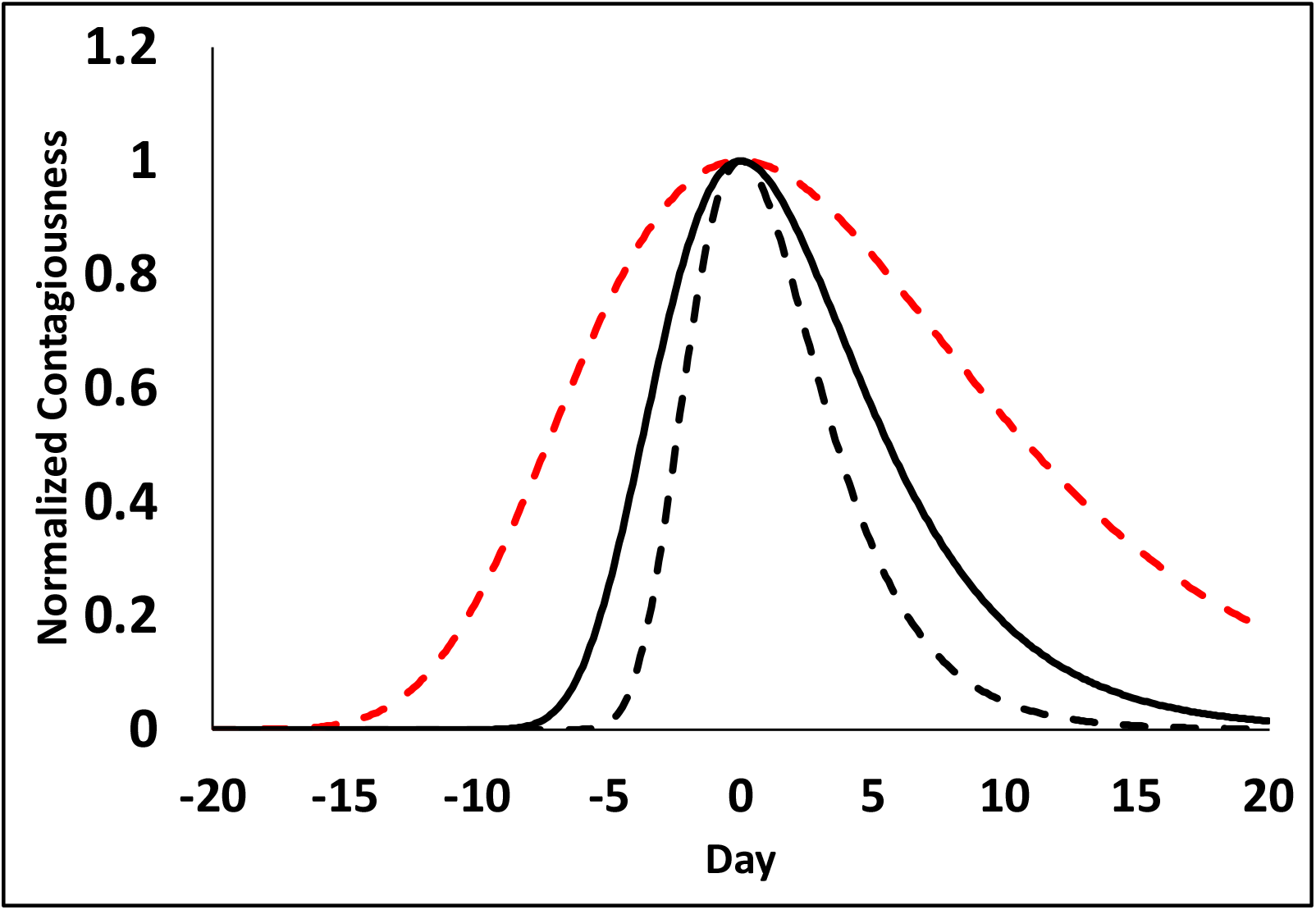
Representation of the normalized contagiousnesss of an initial infected person with one contact with Covid 19. The solid line was generated using the nominal value of *K*_*T*_ = 0.26 in Equation 49; the black dashed line *K*_*T*_ = 0.40; and the red dashed line, *K*_*T*_ = 0.13; the upper and lower 95% confidence intervals, respectively, for *K*_*T*_. The contagiousess begins before time zero because the initial individual was infected before the start of the epidemic.

#### Using the Google Residential Mobility Measure data to Project Country Data

With an estimate of *K*_*T*_ in hand, to project the course of an epidemic using the KMES, we need to also estimate *P*_*c*_. If we first assume that both *K*_*T*_ and *P*_*c*_ are at least piece-wise constant for periods of time, we can differentiate both sides of Equation 28, divide the result by *N*(*t*), rearrange the terms, and then take the natural log of both sides to obtain the following useful expression,

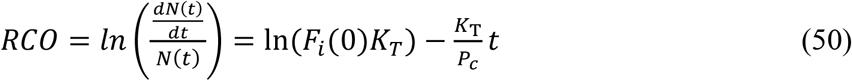

This quantity is labelled the “Rate of Change Operator” (RCO) because it is a measure of the rate of change of *N*(*t*), per person within *N*(*t*). Given estimates for *K*_*T*_ and *F*_*i*_(0), the RCO enables the projection of new cases because the estimation of the current value of *P*_*c*_ can be made from it using only the contemporaneous values of *N*(*t*) and 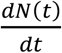.

The process of estimating *P*_*c*_(*t*) begins by first applying Equation 50 to data recorded in six different countries during the initial stages of the COVID-19 pandemic (Roser, et al. 2021), the results of which are plotted in Figure 4. As this figure shows, the RCO curves become straight lines during a period both before and shortly after the date of the imposition of containment actions (arrows in the plots); indicating that *K*_*T*_, *P*_*c*_, and the RCO slope were approximately constant in this period.

**Figure 4.**
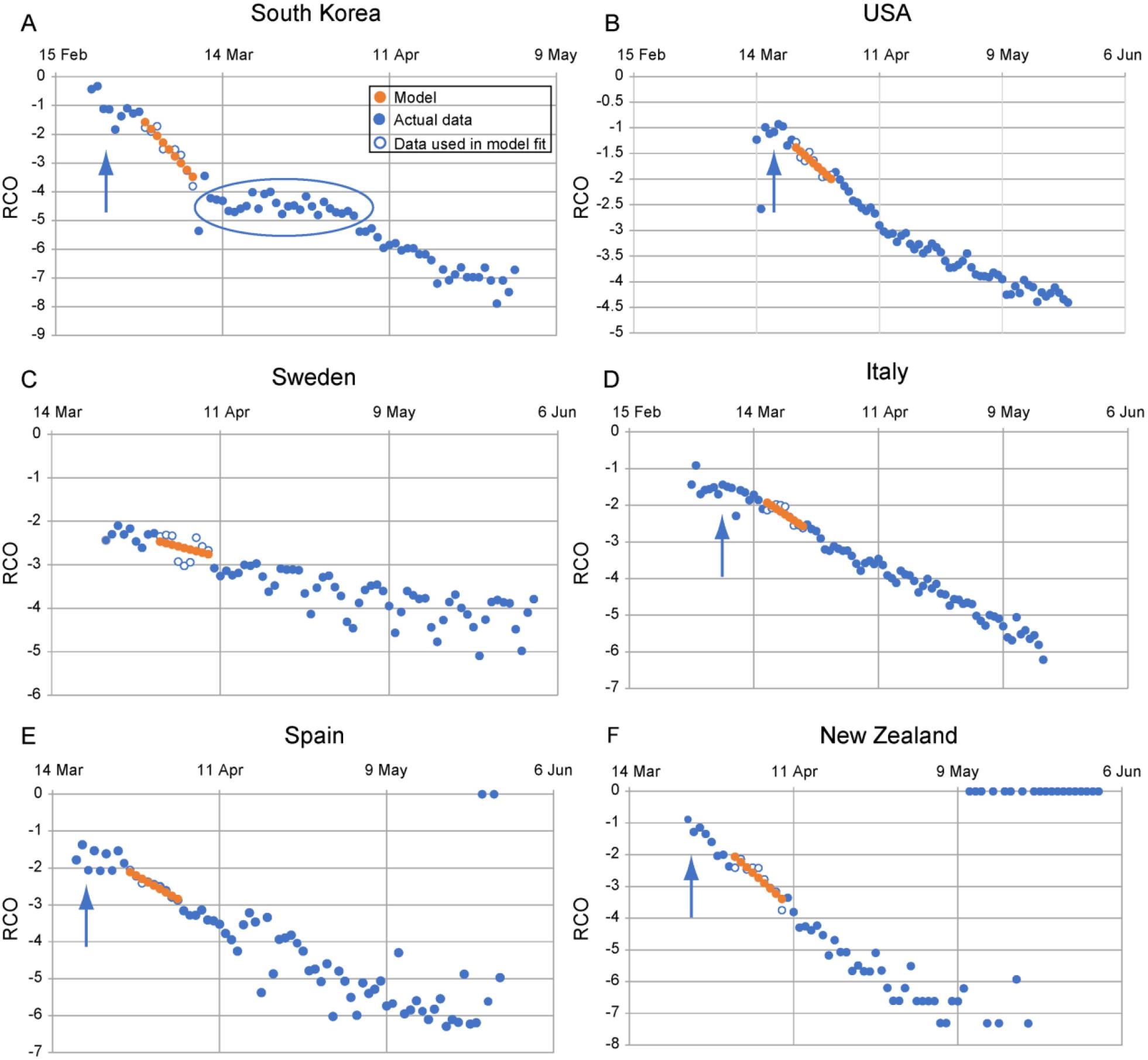
Rate of change operator (RCO) curves for COVID-19 cases in various countries. An epidemic can be described by a piecewise linear model using the RCO (Equation 50). A short segment of orange dots in each graph is a linear fit to the corresponding points (blue/white circles) in the observed data. The slopes of these dotted-line segments are the values of 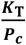 which are tabulated in Table2. In some countries, RCO curves changed markedly soon after the date containment measures were implemented (arrows): **A)** South Korea, February 21; (the oval highlights a departure of the observed data from the RCO slope, indicating failures in, or relaxations of, social distancing); **B)** USA, March 16; **C)** Sweden did not implement any specific containment measures, so the model calibration was begun on April 1, the date when the slope of the RCO curve first became steady. **D)** Italy, March 8; **E)** Spain, March 14; **F)** New Zealand, March 25. All dates are in 2020.

With reasonable confidence that the RCO slope 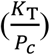 was constant, an initial value of the slope was then estimated by fitting Equation 50 to short, nine data point portions early in the straight segments of each country’s RCO time series. Assuming that this slope was valid for the first date in the series, the value of *P*_*c*_ for each country was next calibrated to Google’s Residential Mobility Measure (Google 2023) on that first date using the expression:

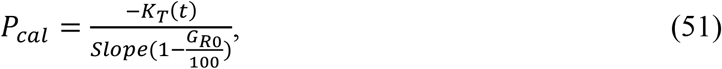

where 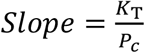 was determined from the early segment of the RCO time series, and *G*_*R*0_ is the value of the Google Residential Mobility Measure on the first date. The *P*_*cal*_ values and dates used for each country are listed in Table 2.

**Table 2.**
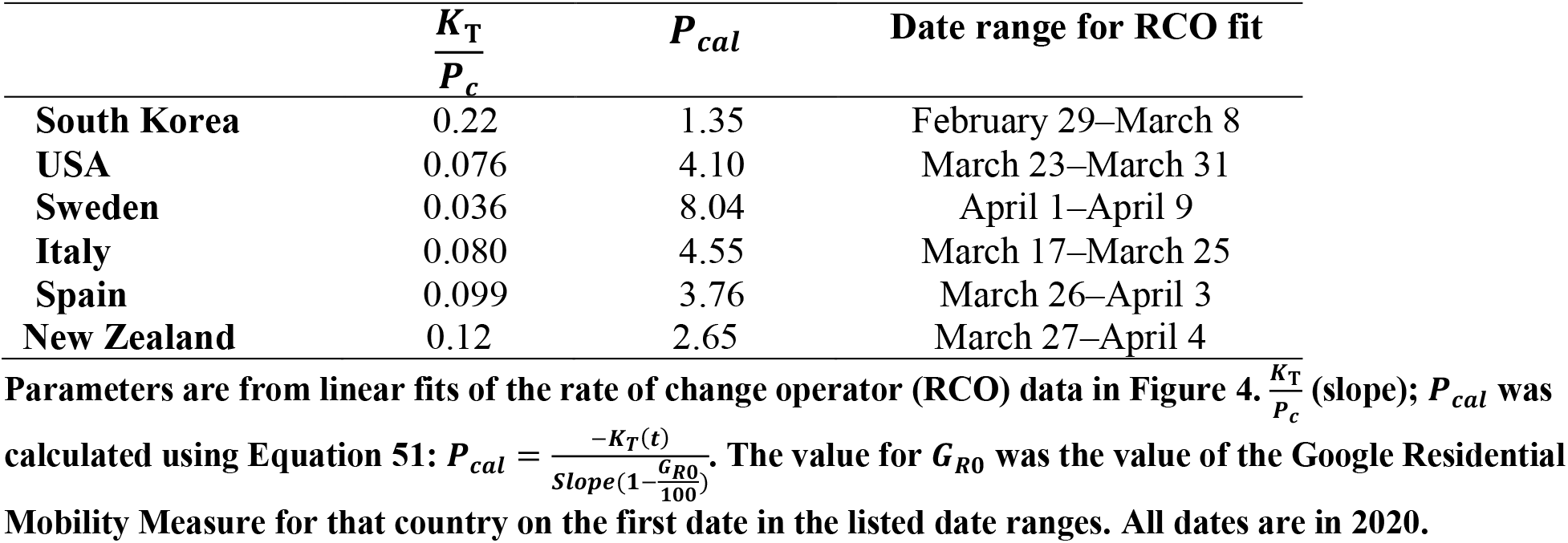
Parameters used to model total and new daily cases of infection for different countries.

Having calibrated *P*_*c*_ to the Google Residential Mobility value on a single date, the subsequent Google Residential Mobility data were then used to find the estimated daily values of *P*_*c*_ by multiplying *P*_*cal*_ and 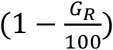 together (where *G*_*R*_ is the Google Residential Mobility Measure for each date). Using the nominal value of *K*_*T*_ from Figure 2 and these daily values of *P*_*c*_, we then employed Equation 6 to plot the daily total cases projections in Figure 5. These projections matched the actual time series with an *R*^*2*^ > 0.85 in each of the six countries for the 60 days following the date *P*_*cal*_ was determined.

**Figure 5.**
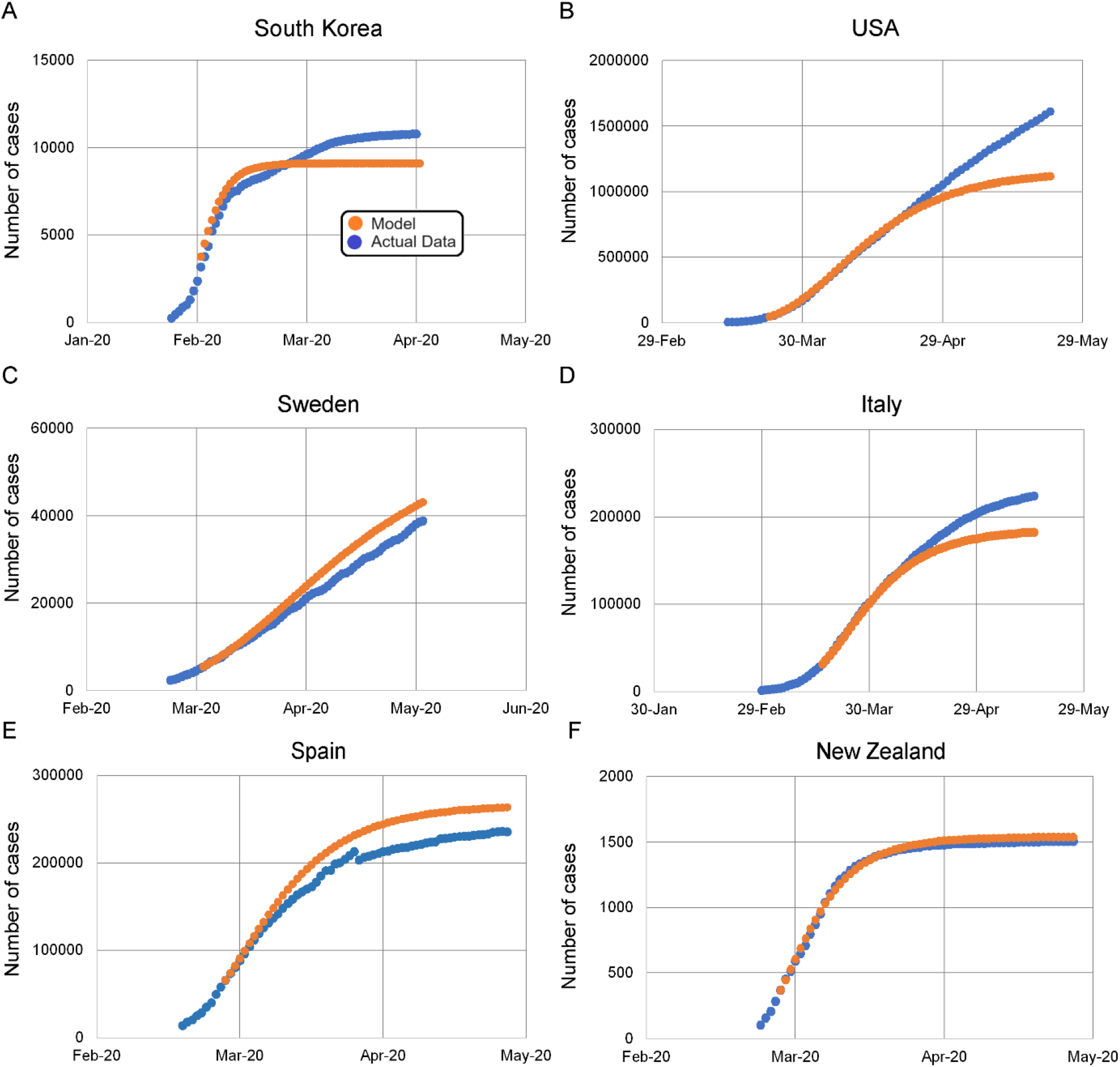
KMES model projections for daily total case counts. **A)** South Korea; **B)** USA; **C)** Sweden; **D)** Italy; **E)** Spain; and **F)** New Zealand. Dots are daily data points observed from (white-center and all blue) or calculated (orange) for each country. The KMES model was calibrated using data from the date ranges listed in Table 2. *R*^2^ > 0.85 for the model fit for all countries for the 60 days after the *P*_*cal*_ was calculated: South Korea, March1-April 29; USA, March 24-May 22; Italy, March 18–May 16; Spain, March 27-Maz 25; New Zealand, March 27-May 25. Sweden April 2-May 31. The deviation of the model from the data in the USA, panel (**B**), after April is elucidated in Supplement 2. All dates are in 2020.

To estimate the values of the daily <u>new</u> cases, Equation 6 was first differentiated to find 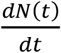, and then, using the previously calculated values of *P*_*c*_(*t*) and *K*_*T*_, the 60-day time series was plotted in Figure 6. These projections have an *R*^*2*^ range of 0.01 (Sweden) to 0.94 (New Zealand); and all projected peaks in new cases were close to the observed peaks in all countries.

**Figure 6.**
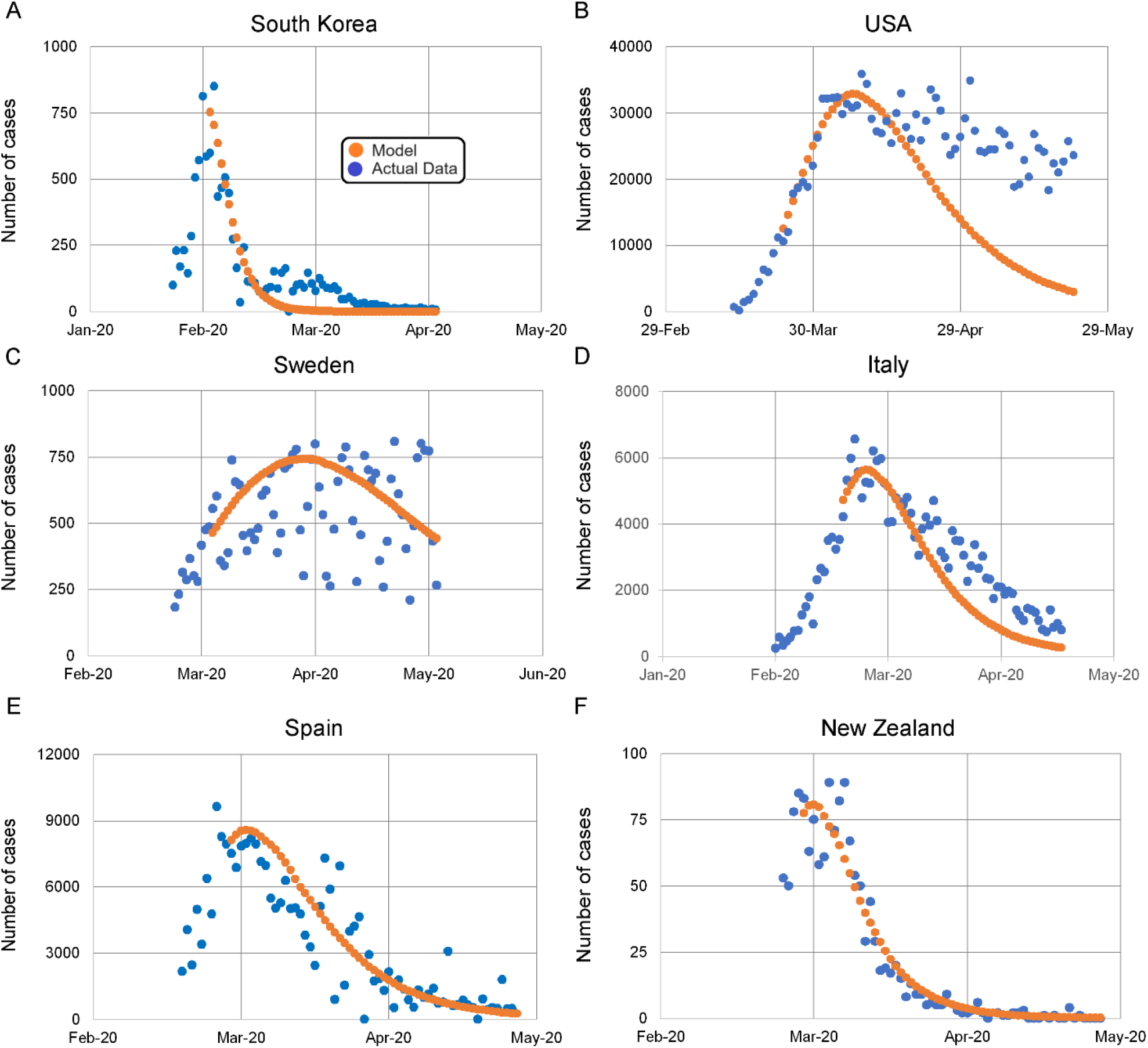
KMES model projections for number of new daily cases. **A)** South Korea, *R*^2^ = 0.87; **B)** USA, *R*^2^ = 0.31; **C)** Sweden, *R*^2^ = 0.01; **D)** Italy, *R*^2^ = 0.88; **E)** Spain, *R*^2^ = 0.82; and **F)** New Zealand, *R*^2^ = 0.94. The orange dotted line is the model in all panels. The blue dots are the daily observations from each country. The. *R*^2^ values are between the model and the data, across countries for the 60 days after the *P*_*cal*_ was calculated.

The projections in Figures 5 and 6 are *<u>not</u>* fits to the full length of the data. Rather, *P*_*cal*_, the prior estimate of *K*_*T*_, and the Google Residential Mobility Measure for each subsequent day were used in the KMES to project the data after the first date.

Since the Google Residential Mobility data produced such accurate projections in these Figures, we conclude that, as we hypothesized in Section 1, that the values of *P*_*c*_(*t*) derived from the model are estimates of the actual average number of infectious contacts that occurred between people during the epidemic. We also comment that the ability of the KMES to project the data so well is a tribute to the ingenuity of Kermack and McKendrick’s model formation.

### Section 3: Managing an Epidemic

An important decision to be made during an epidemic is whether to tighten or loosen restrictions on social interactions. To illustrate how tightening restrictions can affect the epidemic outcome, in Figure 7, plots of the total cases and new cases per day are presented for four countries in two pairs. The pairs were chosen by matching countries with similar population densities, assuming that their baseline social interaction levels were similar. These specific countries were selected, because, in a presumably unintentional natural experiment, the paired countries imposed very different social restrictions at the outset of the COVID-19 pandemic, and as can be seen in the figure, realized very different outcomes.

**Figure 7.**
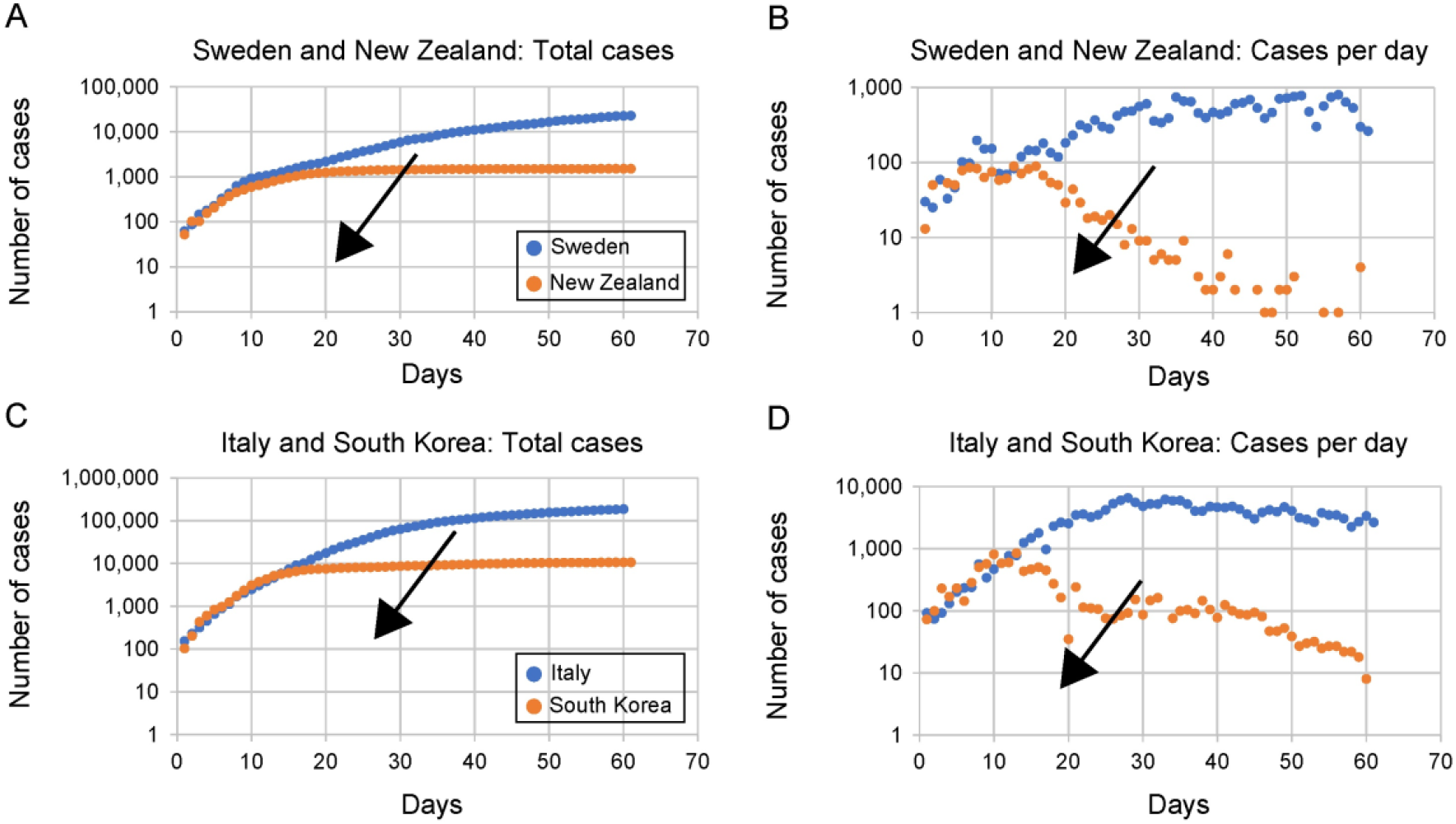
Total cases and new cases per day for countries with comparable population densities, which employed different levels of social containment. The arrows point in the direction the curves trend as the social restrictions were increased. Lower social interaction produced lower total cases and earlier peaks in the new cases per day.

As indicated by the arrows in the plots, in the countries which imposed strict policies, New Zealand and South Korea, the total cases leveled off sooner (Plots A and C) and the new cases per day peaked earlier (Plots B and D) than in their matching countries, Sweden and Spain, which imposed less stringent restrictions.

Of course, it is easy to look back in time at data and prescribe a better course of action, but, since this luxury is not available at the outset of an epidemic, a model that accurately projects trends and provides immediate decision support as the epidemic evolves must be selected. Therefore, to compare models, Equation 28 was used to create a KMES simulation, and a Euler approximation was used to solve the simple SIR model found in Brauer (2008),

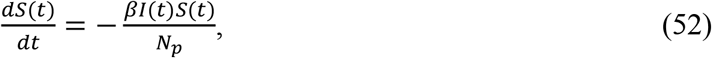

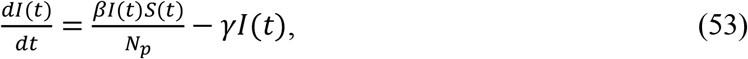

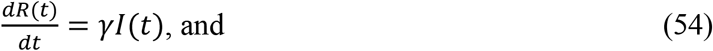

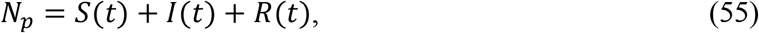

where *β* and *γ* are constants. These simulations were then plotted in Figure 8.

**Figure 8.**
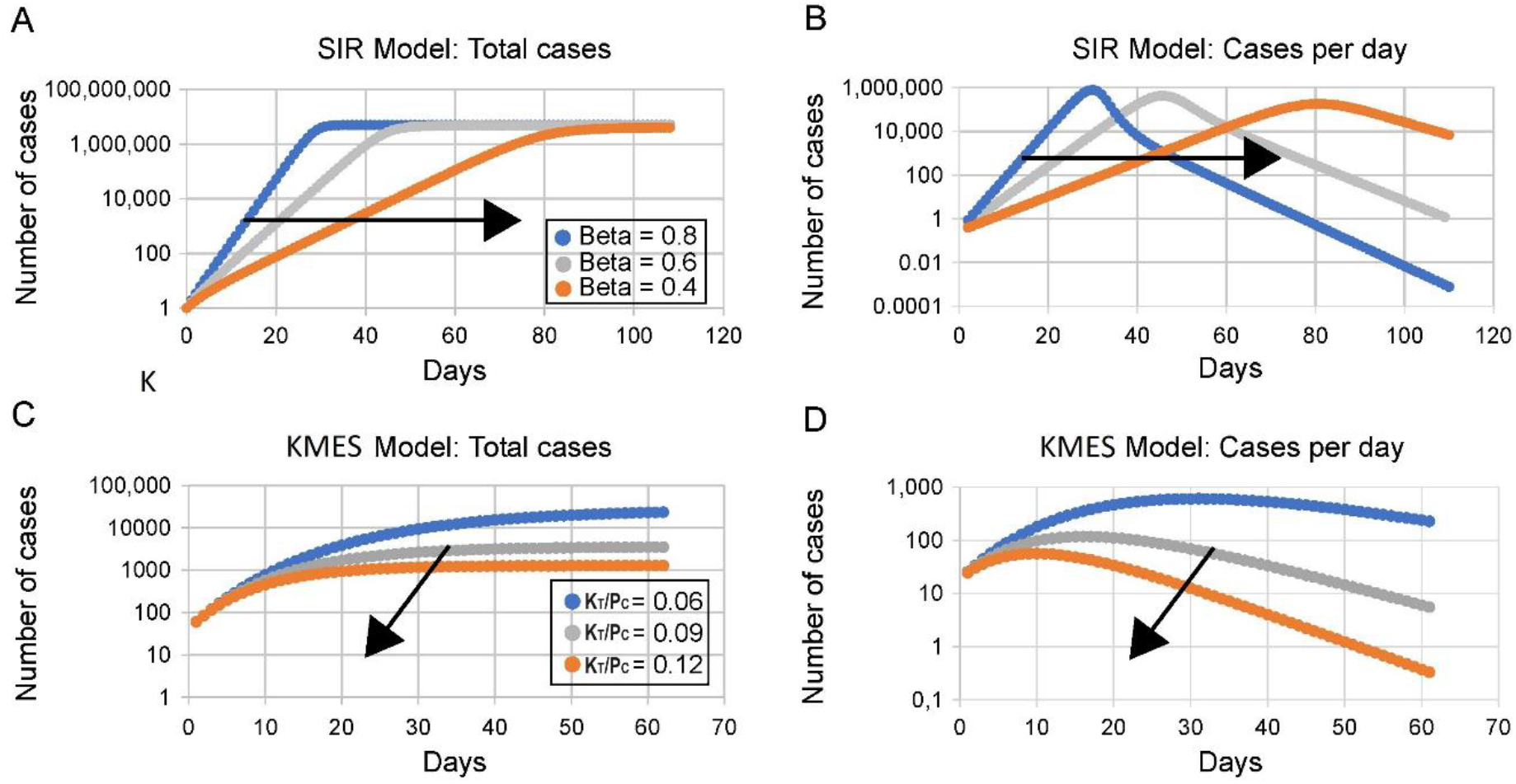
SIR and KMES simulations with varying levels of social restrictions. The arrows point in the direction the curves trend as the social restrictions are increased. The value of *γ* = 0.2 in the upper two panels and *K*_*T*_ = 0.26 in the lower two panels.

As indicated by the arrows in Plots 8A and B, higher social restrictions (lower *β*) in the SIR approximation results in both the plateau of total cases and the peak in new cases occurring <u>later</u>. This illustrates the phenomenon known as “Flatten the Curve” found in both Dilaurio, F., et al (2011) and the analytical results for the SIR model in Kröger and Schlickeiser (2020). However, the trends in plots 8A and B do <u>not</u> match the trends in the country data in Figure 7.

In contrast, as also seen in Figure 8, the KMES projects that a peak in daily new infections will occur <u>earlier</u> (Plot D in Figure 8); and that total infections will plateau sooner and at lower values (Plot C in Figure 8) with higher social restrictions (higher values of *K*_*T*_/*P*_*c*_). These projections match well the trends of the actual case data in Figure 7.

#### Diagnosing the Status of an Epidemic

Given the clear qualitative fidelity of the KMES projections to the actual trends in the country data, we now consider the ability of the KMES to accurately project future case numbers beyond the time frame shown in Figures 5 and 6, and without the daily knowledge of the Google Residential Mobility values. We do this using the 7-day average of United States total cases data (Roser et al, 2021) from the COVID-19 pandemic over an 18-month period.

The total cases data were projected by first calculating the value of the RCO on a given day and the eight previous days. Equation 50 was then fit to those nine known RCO values to find their intercept, ln(*F*_*i*_(0)*K*_*T*_), and slope, 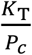. These parameters were assumed to be constant throughout those prior eight days and the following 21 days, a total of 30 days. They were next used in Equation 28 with the time set to 21 days, along with the value of the total cases on the chosen day, to project the total cases 21 days into the future for each date. Figure 9 is a plot of that projection overlaid onto a plot of the actual 7-day average total case data for 18 months following April 30, 2020. As seen in the figure, the KMES projected the total cases 21 days in advance with a Mean Absolute Percentage Error of 4.1% over the entire period.

**Figure 9.**
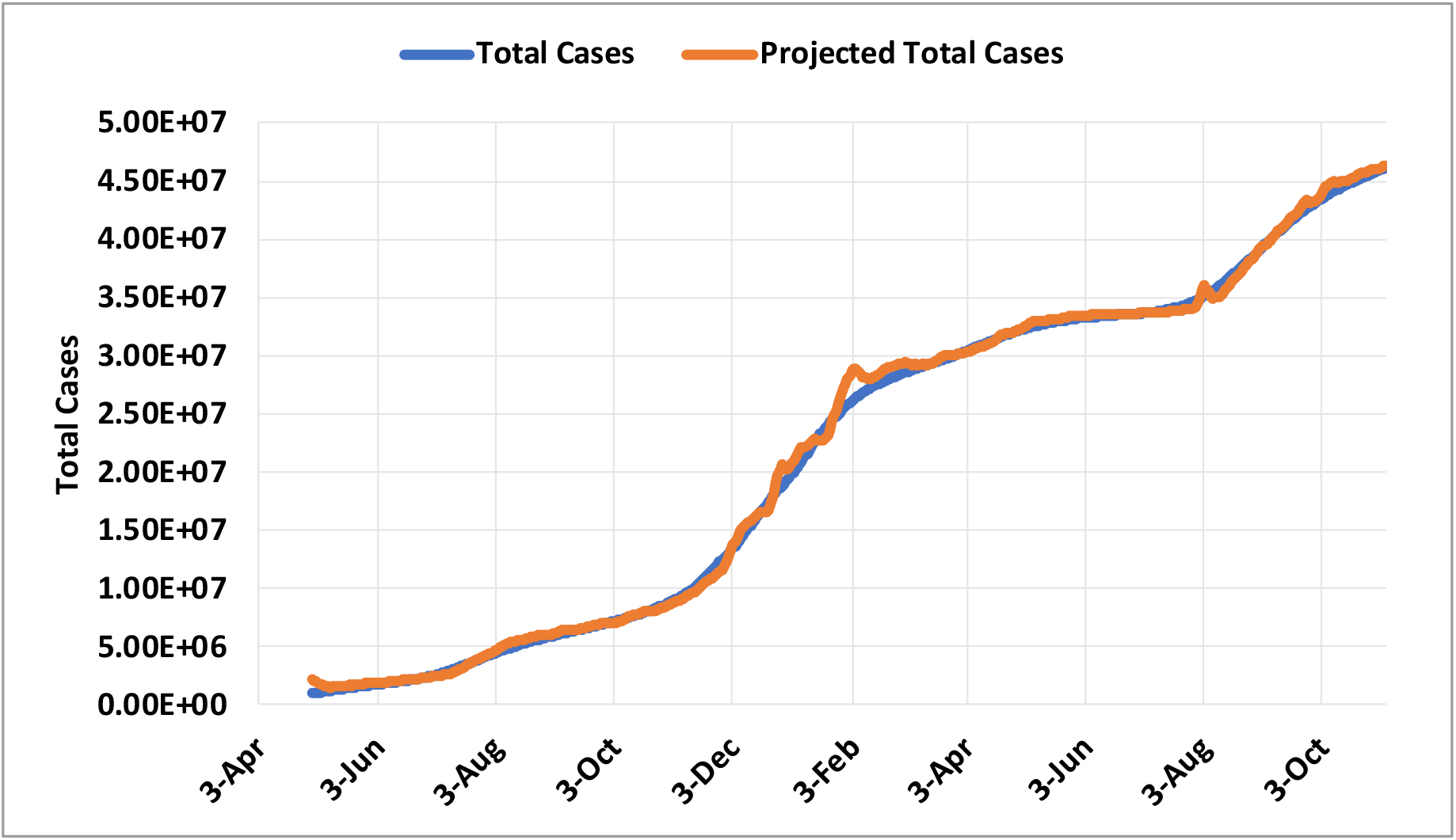
Projection of cases in the United States from April 25, 2020 to November 4, 2021. The Orange line is the projection of total cases 21 days in the future from the point of each actual data point. The total cases data points are the average of the 7 prior days of total cases. The data ranges from April 30, 2020 to November 4, 2021.

Although Figure 9 demonstrates that the KMES can be used to project the potential daily new cases, and, presumably, the hospitalizations, three weeks in advance, a projection alone cannot achieve another paramount goal of public health management: recognizing and avoiding outbreaks. Fortunately, as a complement to projections, the *R*_*Eff*_ calculation can be used to detect emerging outbreaks, because it reflects the underlying dynamics, anticipates the future, and, its daily value can always be calculated using this restatement of Equation 31,

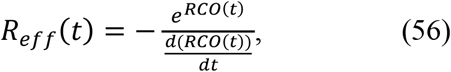

If we discretize Equation 56 and use the relationship: 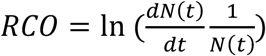, we obtain this formula for *R*_*Eff*_,

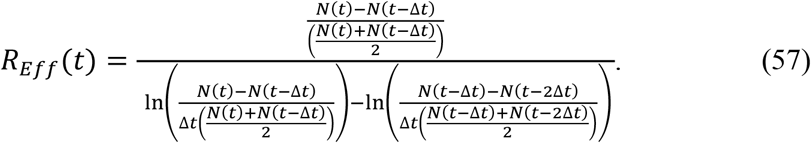

As Equation 57 illustrates, the determination of whether the epidemic is growing or declining, can be made by using just the most recent <u>three known</u> consecutive values of *N*(*t*).

When using Equation 57, if an outbreak occurs, the average number of susceptible people within the *P*_*c*_ groups has risen at a fast rate. The fraction infected will have also risen because the contagious people newly in contact with the susceptible people in the outbreak will have become more infectious due to these new contacts. This increase in the fraction infected, if it is large enough, can cause the slope of the RCO to become positive because it also effectively causes the value of *F*_*i*_(0) in Equation 50 to become larger. This mathematical effect, if present in the data, will cause the value of *R*_*Eff*_(*t*) to be less than zero, an obviously unphysical result. This mathematical artifact occurs because we derived Equation 50 under the assumption that *P*_*c*_ is a constant.

Pragmatically, since this artifact is known to only appear when the outbreak is large enough, whenever Equation 57 returns a negative value for *R*_*Eff*_(*t*), these times can be deemed to be outbreaks. Therefore, when using Equation 57, the outbreaks should be identified as time intervals when Equation 57 returns a value *R*_*Eff*_(*t*) < 0 or *R*_*Eff*_(*t*) > 1.

To demonstrate the use of Equation 57, we first restated the data that was used in Figure 9 by aggregating the total new cases 7 days at a time. We used 7-day aggregates because often epidemic data is only available in this form and because a 7-day aggregation smooths the variations in daily case counts. We then applied Equation 57 to this data and plotted the result in Figure 10A. Accordingly, the pink shaded areas indicating the outbreaks in Figure 10 are those times when Equation 57 returned a value *R*_*Eff*_(*t*) < 0 or *R*_*Eff*_(*t*) > 1.

**Figure 10.**
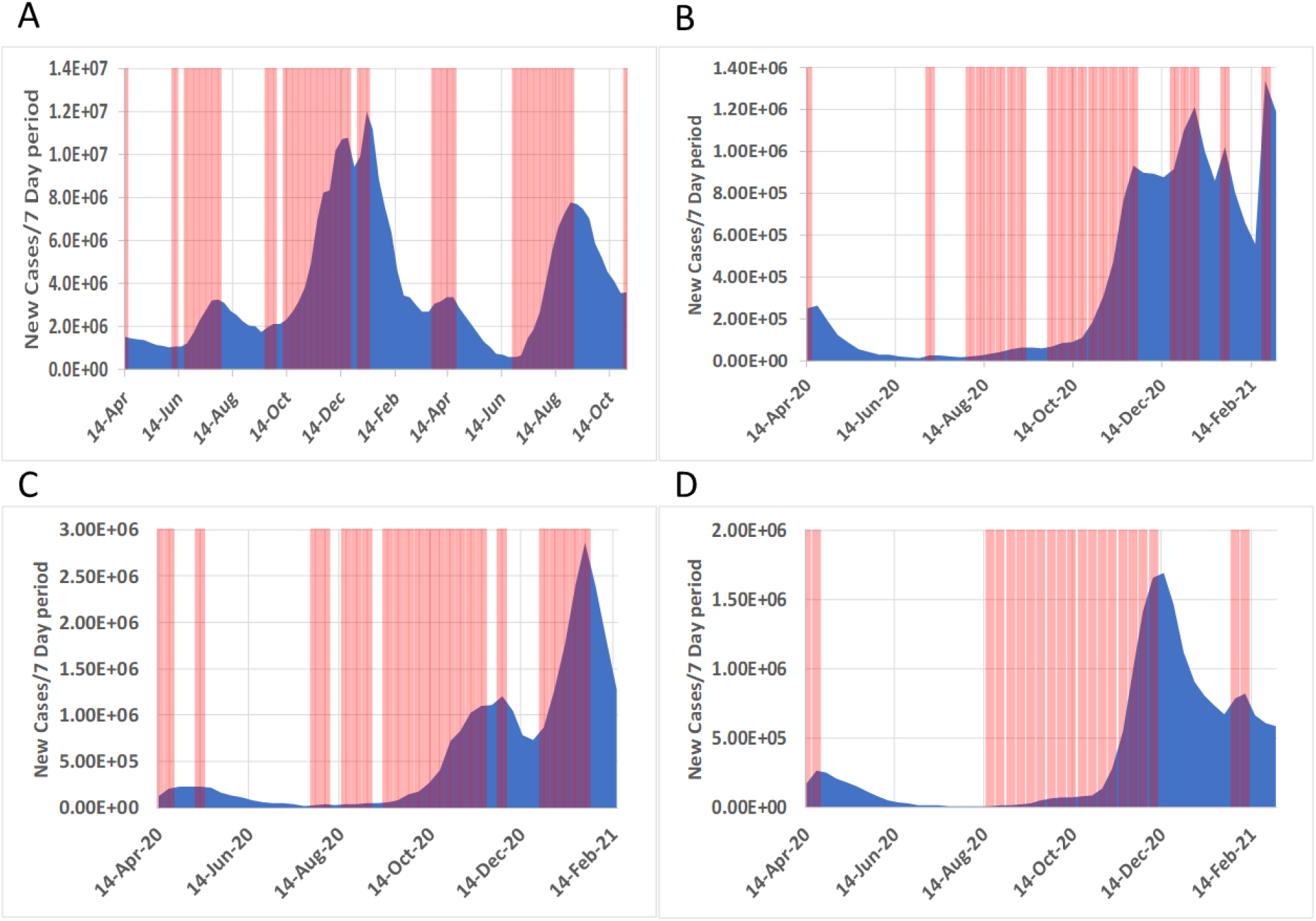
Periods when *R*_*Eff*_ indicates an outbreak or peak. The blue area indicates the 7-day total new cases for the dates indicated. The pink shaded areas indicate the periods when the *R*_*Eff*_ was above 1 or below 0. The explanation of why *R*_*Eff*_ can go below 0 is explained in the text. A; United States April 14, 2020 to November 2, 2021; B: April 14, 2020 to March 2, 2021; C: United Kingdom April 14, 2020 to February 16, 2021; D Italy April 14, 2020 to March 2, 2021.

As expected, the pink shaded time periods in the figure mark the periods of the outbreaks very closely. Panels B, C, and D in Figure 10 are additional plots created using the same procedure for case data from Germany, the United Kingdom and Italy respectively. These additional plots show the same correspondence between the pink areas and the outbreaks for the dates covered. Figure 10 demonstrates that Equation 57 could be a powerful tool for use in determining if the population restrictions are strong enough to prevent major outbreaks.

There are no uncertainty intervals around the locations of the pink shaded areas because their position in time was dependent solely on the differences between two consecutive data points of total cases without the use of fitted parameters. The only errors present in the calculation are associated with measurement errors of *N*(*t*) which were not estimated. The calculation is right censored because data from the past and present were used to determine the position of the pink shaded areas.

#### Controlling the Epidemic

If the estimation of *R*_*Eff*_ identifies an outbreak, social policies should immediately be modified to blunt the impact. In this scenario, expressions based on the KMES can be used to set out a program for gaining control of the emerging outbreak in a fashion consistent with social and political realities.

Differentiating Equation 28 twice produces the epidemic acceleration,

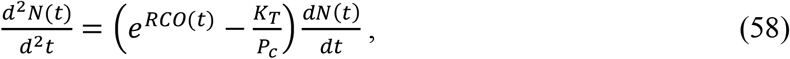

which can be used to quickly determine whether the control measures in place, embodied by *P*_*c*_, are sufficiently effective. When the term, 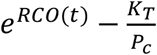, on the right side of Equation 58 is positive, then the control measures are not strong enough; and conversely, when this term is negative, the epidemic is being brought under control, a condition coincident with 0 < *R*_*Eff*_ < 1.

The maximum value of *P*_*c*_ that will begin to bring down the new cases per day is the quantity upon which management strategies pivot. By setting the left-hand side of Equation 58 to zero, then solving for *P*_*c*_, a defining relationship for this critical objective is found:

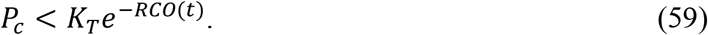

Equation 59 succinctly states that *P*_*c*_ must be managed to stay below 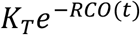 to ensure that the acceleration is negative and therefore, the epidemic will slow. Since the *RCO*(*t*) can be computed every day and *K*_*T*_ can be estimated using the technique illustrated in Section 2, the maximum level of infectable social contact allowable (*P*_*c*_ in Equation 54) to start or continue decreasing the number of new cases per day can always be calculated. As a side comment, and as already explained, if the slope of the RCO curve is determined from the graphical analysis to be greater than zero, then an outbreak has occurred, *P*_*c*_ can be assumed to be too large, and immediate reductions in social interactions, based on the following procedure, are needed.

If the situation warrants, the time needed to reach a desired reduction in 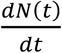, at a future time, *t* + *t*_*target*_, can be calculated for a given level of social interaction by first defining the desired fractional reduction in cases as *D*_*tf*_,

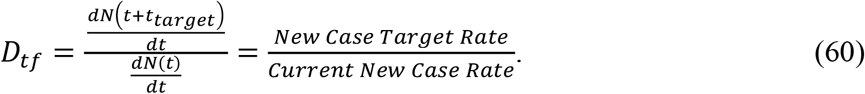

Using the derivative of Equation 28, the following expression emerges,

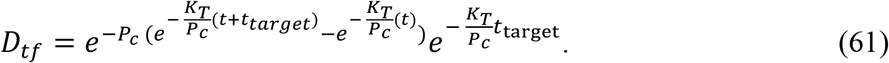

If *t* ≫ *t*_*target*_, then 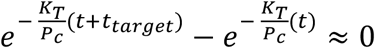, and the following can be deduced from Equation 61,

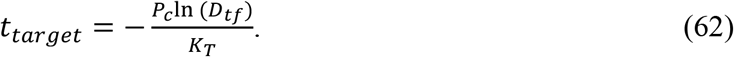

Equation 62 quantitates the number of days, *t*_*target*_, that a level of social containment, *P*_*c*_, must be maintained to reach a desired reduction, *D*_*tf*_, of the current daily cases.

As an example, in the case of the United States, based on the data shown in Figure 10, a very large outbreak started during the last days of September and early October 2020. On October 10, 2020, there were 58,082 new cases and the outbreak peaked 90 days later with 283,204 recorded new cases. Had the US implemented a social program to reduce the number of average infectious contacts (*P*_*c*_) to 10 people for those same 90 days, instead of an outbreak and a peak, Equation 62 projects that the number of new cases on January 8, 2021 would have been approximately 5800, a 98% reduction from the actual value. In Supplement 2, we present further examples and additional insights utilizing the data in several additional countries.

## Discussion

In response to Diekmann’s call for action, the “wisdom” we have found in the Kermack and McKendrick integro-differential equations appears to be substantial. Foremost, we suggest that the KMES obviates most of the need to use any approximation to the integro-differential equations. Secondly, the KMES appears to accurately project the dynamics of an actual pandemic as seen in multiple independent data sources and it offers a pathway to new analytical expressions useful in characterizing and managing an epidemic.

The hybrid path between network concepts and deterministic equations that we used to derive the KMES also suggests that there is potential to develop more sophisticated hybrids. Further exploration of the means to specify *P*_*c*_(*t*) within network types that are representative of physical interactions between people may well lead to still more accurate forecasts of epidemic progressions. Such extensions could be applied to influenza epidemic data and data from a myriad of other diseases, including the next SARS virus redux. Hybrid approaches such as these could also reduce the computational burden associated with models that employ extremely high numbers of network nodes by allowing the calculation of results for portions of the network using analytical expressions rather than simulations.

The analysis can also be further improved by incorporating the enormous amount of case data now available from the COVID-19 pandemic. This additional analysis can improve the estimate of the key parameter, *K*_*T*_(*t*), including variations in time (with mutations of the infectious agent) and possibly with local genetic variations in the population affected. This could improve the basis for establishing the actions people and governments need to take to achieve target values of *P*_*c*_(*t*).

Although we highlighted that a disease with a latent period would manifest itself by affecting the value of *K*_*T*_(*t*), we did not explore the analytical consequences. The model can be further developed by definitively separating the population of inactive or latent infections from the contagious infections within *I*(*t*) and then exploring the effect of the latent period on the solution parameters. The resulting analyses may establish whether the latent period can be determined during the early epidemic or possibly be ignored once the epidemic has fully developed. Both would be useful outcomes.

With regards to the KMES structure, it is reassuring that it has the intuitive form, encapsulated in Equation 25:

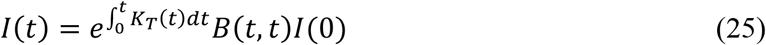

This equation states that the input of infections, *B*(*t, t*)*I*(0), is transformed into the time varying output of infectiousness, *I*(*t*), through an exponential step response function, 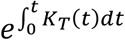. Our analysis has led us to affirm this obvious-in-retrospect, practical mathematical characterization of epidemic dynamics. It is additionally reassuring that bridges, such as in the final size expressions and the basic reproduction number, can be built between the KMES and the SIR approximations.

The KMES also meets other expectations. For example, we recognize Equations 6 and 28 as Gompertz equations. This form is highlighted in the analysis of Onishi et al (2021) who demonstrated that the COVID-19 epidemic time course in many countries was well fit by a Gompertz model. These authors did not offer a basic principles argument as to why this is so, but they found and report a strong correlation to this aspect of our model structure. Additionally, using the independently measured population mobility in the Google data, we found that the KMES accurately projects phenomena which arose in the COVID epidemic. This also confirms the intuition that this independently sourced data on the movement of populations should be useful in projecting epidemic dynamics.

Our expressions for the time to the peak fit well the data from different countries which imposed very different containment strategies. These equations show that as people interact less frequently (social containment is increased), the peak number of infections is much lower, and occurs earlier. The KMES shows that strong containment actions shorten an epidemic, as data from several countries clearly demonstrate; and as one would intuit.

Lastly, the discovery of a very simple method to assess whether an outbreak is occurring, embodied in *R*_*eff*_ offers the promise of a new quantitative tool to guide public health officials in their decision making. We also hope that our work here serves as an impulse for the reinvigoration of epidemic modelling using the core of Kermack and McKendrick’s original work.

## Data Availability

All data produced in the present study are available upon reasonable request to the authors

## Abbreviations used in this text

KMES: Kermack McKendrick integro-differential Equation Solution
RCO: Rate of Change Operator
SIR: Susceptible–Infectious–Recovered
CI: Confidence Interval

## Supplement 1: Solution to the Kermack and McKendrick Equations

The path to the KMES is complex, therefore, we provide the following short guide:

1. First rewrite Equations 1 through 5 in terms of time alone utilizing the weighted averages of functions of *φ*(*t, θ*) and *ψ*(*t, θ*).
2. The general solution to these rewritten equations is developed, and the relationships between key parameters are clarified.
3. Network model concepts are used to derive an expression for the effect of the population interactions on the evolution of the term 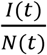, the fraction of those ever infected, that are currently infectious.
4. This expression opens the pathway to the final form of the KMES which is expressed in terms of the parameters *K*_*T*_(*t*) and *P*_*c*_(*t*), the disease transmissibility and the population interactions, respectively.

### Step 1: Rewrite the Integro-Differential Equations

As a reminder, the Kermack and McKendrick equations are:

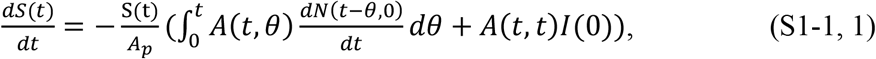

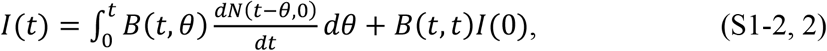

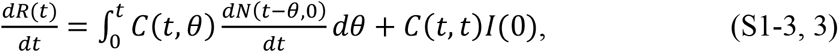

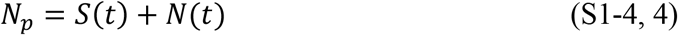

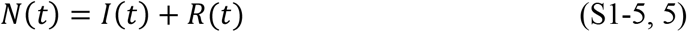

where 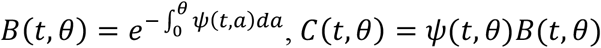, and *A*(*t, θ*) = *φ*(*t, θ*)*B*(*t, θ*). *N*_*P*_ is the total population, *A*_*P*_ is the area that contains *N*_*P*_, *θ* is the time since infection of any member *N*(*t*), and 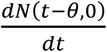 equals the new infections at time *t* − *θ*.

Since *A*(*t, θ*) = *φ*(*t, θ*)*B*(*t, θ*), the weighted average of *φ*(*t, θ*) at a time t is,

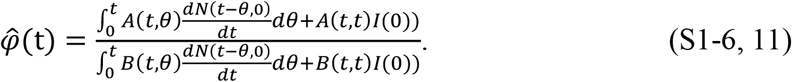

Likewise, since *C*(*t, θ*) = *ψ*(*t, θ*)*B*(*t, θ*), the weighted average of *ψ*(*t, θ*) at time t is,

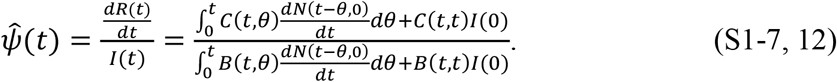

It is also useful to define a parameter *K*_*T*_(*t*) as,

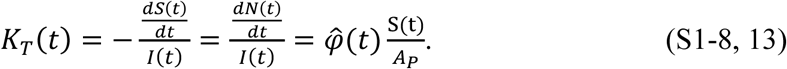

Using these definitions, Equations S1-1 through S1-5 can be rewritten in terms of time as,

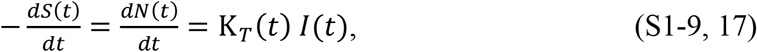

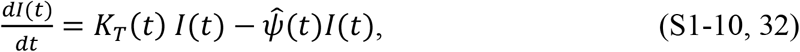

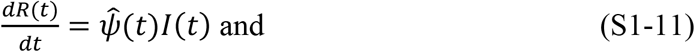

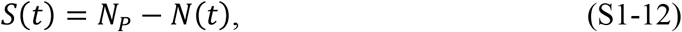

where *N*(*t*) = *I*(*t*) + *R*(*t*).

### Step 2. Solve the Rewritten Equations

The solutions to Equations S1-9 and S1-10 are,

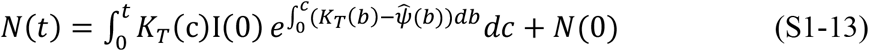

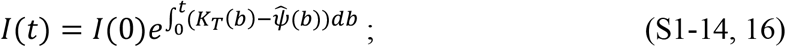

and, since *R*(*t*) = *N*(*t*) − *I*(*t*), Equation S1-13 and S1-14, when combined, solve Equation S1-11.

Equations S1-13 and S1-14 solve Equations S1-9 to S1-11, but since they are written in terms of 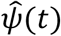 and *K*_*T*_(*t*), neither of which appear in Equations S-1 through S1-5, it is unclear whether they also solve Equations S-1 through S1-5. Therefore, we will now explain in detail how 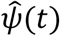 and *K*_*T*_(*t*) can be expressed in terms of *ψ*(*t, θ*) and *φ*(*t, θ*) to ensure that Equations S1-13 and S1-14 also solve Equations S1-1 through S1-5.

We begin by rewriting Equation S1-7 as,

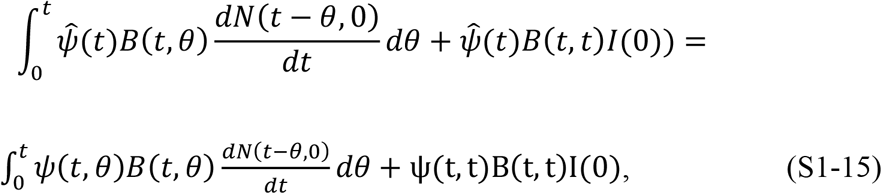

which can be simplified further using the definition of *I*(*t, θ*) and Equation S1-8,

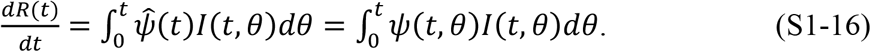

Since 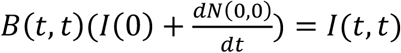, both 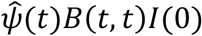 and *ψ*(*t, t*)*B*(*t, t*)*I*(0) in Equation S1-15 are included in the integrals in Equation S1-16.

Equation S1-16 states that we can assume 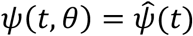 for all *θ* and *t* without affecting either the value of the integrals in Equation 16 or the value of 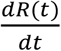. Because we are only interested in the sum of the change in the number of infectious people at any chosen time, the actual values of the individual *ψ*(*θ*) terms are irrelevant when evaluating the integral in S1-16; and, therefore, we adopt the assumption that 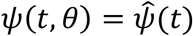 for the remainder of the analysis.

Using Equation S1-16 and the equivalency 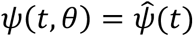, we can now derive *B*(*t, θ*) in terms of 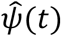 to assist us in proving the solution. We begin by noting that Kermack and McKendrick created the function, 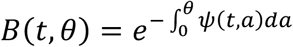, using the right-hand side integrand in Equation S1-16 to explain that, in each time step, every θ-group (*I*(*t, θ*)) recovered by a proportion *ψ*(*t, θ*). We can rederive their formula by starting with the derivative of the infectious expressed as,

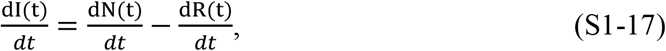

which can be discretized to,

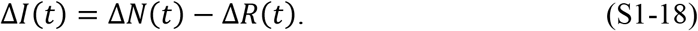

Equation S1-18 can be rewritten using the discrete forms of both Equation S1-16, the definition 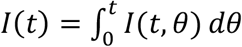, and keeping in mind that Δ*t* = Δ*θ*, as,

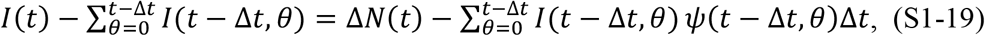

and then as,

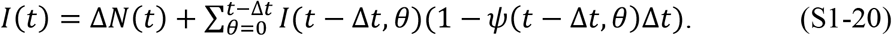

The right-hand side of Equation S1-20 can be expanded by an additional term and written as,

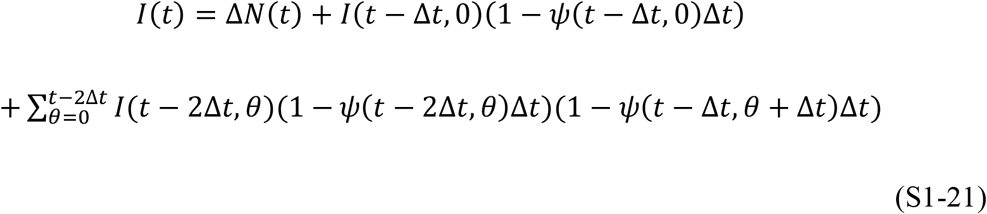

This process can be repeated by continually writing out another term on the righthand side until the following sequence of terms appears,

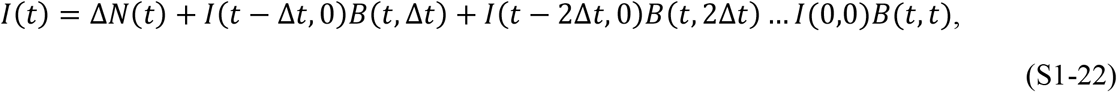

where,

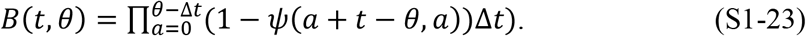

We comment here that Kermack and McKendrick assumed that *ψ* was only dependent on *θ* and not on time. Although this is unnecessarily restrictive from both a mathematical and real-life standpoint, applying their assumption to the expression for *B*(*t, θ*) (Equation S1-23) and then taking the limit as Δ*t* goes to zero, we have, 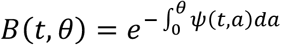; Kermack and McKendrick’s expression for *B*(*t, θ*).

Since we have already demonstrated that we can assume 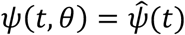 for all *θ* without affecting the value of the integral in Equation S1-16, and the right most summation that we used in Equation S1-19 is the discrete form of this integral, we can replace *ψ*(*t, θ*) with 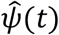 in Equation S1-19 and repeat the sequence of operations from Equation S1-19 through S1-22. Written in detail, the resulting product sequence for *B*(*t, θ*) is,

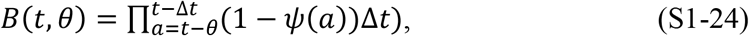

which, in the limit as Δ*t* → 0, becomes 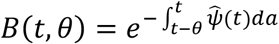, the relationship we seek. This important result also shows that when 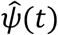 is used, the integral limits in *B*(*t, θ*) must be the interval from *t* − *θ* to t, the same time interval covered within Equation S1-23 as Δ*t* → 0. The use of this interval also ensures we use the proper values of 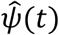 in the restated function, *B*(*t, θ*) Proceeding with the proof of the solution, we note that because 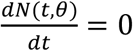 for all *θ* > 0, then 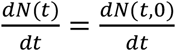. We can use this relationship, substitute Equations S1-14 and S1-9, along with the expression 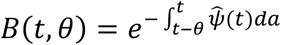 into Equation S1-2; and, keeping in mind that *dθ* = *dt*, rewrite Equation S1-2 as,

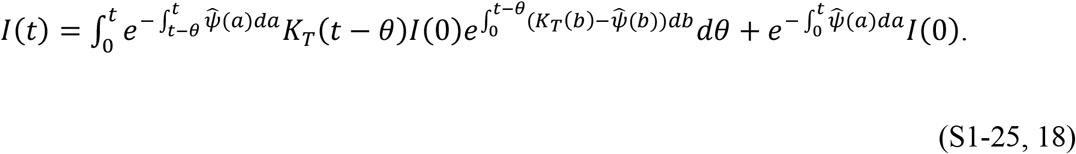

Equation S1-25 can be simplified to,

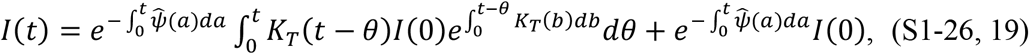

and since 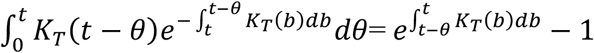, Equation S1-26 reduces to:

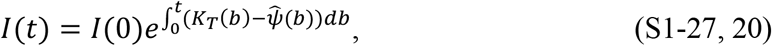

which is identical to Equation S1-17, thereby proving that Equation S1-14 is a solution to Equation S1-2.

We can now prove that Equation S1-13 is a solution to Equation S1-1 in a similar manner. The first step is to recall that 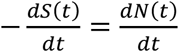 and then differentiate both sides of Equation S1-13 to find, 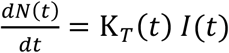 which is Equation S1-9. We then substitute the expression 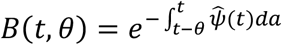 along with Equations S1-9, S1-14, and S1-6 into Equation S1-1 to obtain,

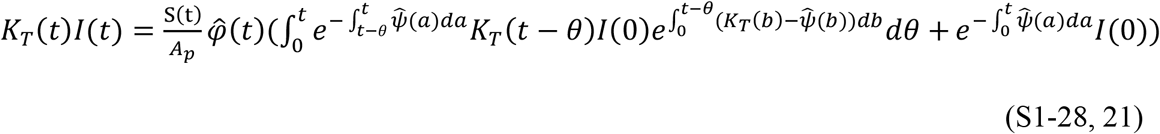

Equation S1-28 can be simplified using Equations S1-8, S1-26, and S1-27, to obtain,

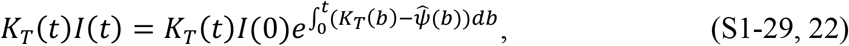

If we then substitute Equation S1-9 into S1-29 and integrate, we find the following expression,

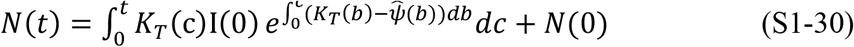

which is identical to Equation S1-13 proving Equation S1-13 is a solution to Equation S1-1.

We also note that by dividing *N*(*t*) into Equation S1-9, integrating, and exponentiating, *N*(*t*) can be restated in terms of 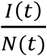 as,

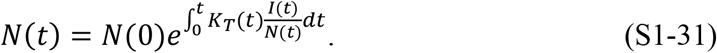

This expression is useful in the next two steps of the derivation.

### Step 3: Include the Effect of the Population Interactions

In their development of a pragmatic model to characterize epidemics, Kermack and McKendrick assumed that the disease transmissibility, together with the population behavior in the guise of density, were the controlling factors for epidemic outbreaks. We note here that we have already proposed *K*_*T*_(*t*) as the disease transmissibility, and the use of *K*_*T*_(*t*) in Equation S1-9 removes the effect of the population behavior from the solution to our rewritten equations. Therefore, in this step of our derivation, by combining Kermack and McKendrick’s concepts with those of network theory, we reintroduce a parameter of population behavior to the solution in Equations S1-13 and S1-14.

We first assume that the population is a network with individuals as vertices and edges capable of transmitting an infection. As a mental image of a spreading epidemic, we further assume that within this network, there is a subnetwork wherein only those contacts that already have, or eventually will, pass on an infection are its edges. We also assume that, in this subnetwork, the mean number of these edges per person, which we call *P*_*c*_(*t*), is the self-weighted mean degree of the subnetwork minus 1. We subtract 1 because a person cannot infect their infector. As defined, *P*_*c*_(*t*) is equivalent to the basic replication number, *R*_0_, as it is known in conventional epidemiological modelling terminology.

In this concept, as the contacts between members of the population change, *P*_*c*_(*t*) can hypothetically vary in time to take any value between 0 and *N*_*P*_. However, since people can only interact infectiously with local persons, it is implausible that *P*_*c*_(*t*) would ever attain a value approaching *N*_*P*_. Therefore, in practice, *P*_*c*_(*t*) is virtually certain to be a number much less than *N*_*P*_. Whereas Kermack and McKendrick assumed that all susceptible people are continually in contact with all infected people (the well-mixed assumption), our concept of *P*_*c*_(*t*) is a more sensible and possibly more general casting of their contact concept.

Kermack and McKendrick also continually reduced the contacts within their model as the susceptible population decreased because they multiplied the number of susceptible people by the number of infected people, a consequence that the use of the parameter *K*_*T*_(*t*) obviates. In contrast, as a more realistic alternative, we assume that the value of *P*_*c*_(*t*) does <u>not</u> change when an infected person infects another person. The disease state of the population within *P*_*c*_(*t*) may change, but *P*_*c*_(*t*)’s value can only change if the behavior of the population changes.

By expressing Kermack and McKendrick’s population interaction notions using a network model, we also interpret their concept of population density as meaning the number of infection-passing contacts between people, which in our parlance is *P*_*c*_(*t*). Within this network, in which density is a proxy for social interaction, individuals can increase or decrease their interaction by creating or breaking contacts (or edges in network terminology), and therefore change the degree between themselves and local people. Emphasizing “local”, the use of *P*_*c*_(*t*) recognizes that the contacts and therefore infections can only occur between people who come to be adjacent to each other. Also, in this network, other than changes due to the addition of outside people or changes in social interaction, in accordance with our first thought experiment, we assume that everyone remains durably in contact with their initial contacts.

With these concepts in mind, we mathematically define *P*_*c*_(*t*) as,

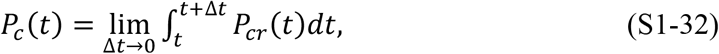

where *P*_*cr*_(*t*) is the average rate of prior or eventually successful infection transmitting contacts for the entire population. Consequently, *P*_*c*_(*t*) is understood to be the instantaneous average number of eventually successful infectious contacts within the population. These contacts are allowed to be fractions of a whole between any two contacts; therefore, there is an amplitude associated with every contact, and it is the average amplitude of these contacts per individual that equates to *P*_*c*_(*t*) for the population.

If, in addition, we define *P*_*S*_(*t*) as the <u>average</u> number of remaining susceptible people in each *P*_*c*_(*t*) group that people in *N*(*t*) will infect, we can deduce a useful relationship between *N*(*t*), *I*(*t*), *P*_*c*_(*t*), and *P*_*S*_(*t*). Because the people in *I*(*t*) are only infectious to the extent that they contact people who are susceptible to the disease agent, by definition, *I*(*t*)*P*_*c*_(*t*) is the total number of current contacts within *N*(*t*) that will eventually be successful. Thus,

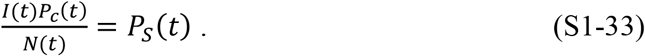

and,

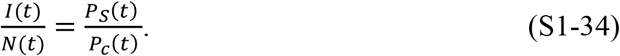

From this insight we see that, while the sizes of each *P*_*c*_(*t*) are uniform, the quantities of susceptible people within individual *P*_*c*_(*t*) groups are not the same.

Symmetrically, since, by definition, *R*(*t*)*P*_*c*_(*t*) is the number of noninfectious contacts within *N*(*t*), if *P*_*I*_(*t*) is defined as the average of previously infected people in the *P*_*c*_(*t*) groups that people in *N*(*t*) <u>cannot</u> infect, then,

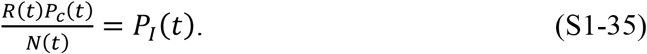

To verify that the definitions are complete and not overlapping, we multiply both Equation S1-33 and Equation S1-35 by *N*(*t*), and then add the result, to obtain the identity,

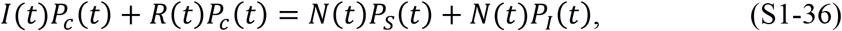

thereby showing that both sides are equal to *N*(*t*)*P*_*c*_(*t*), and there is no overlap.

The next step is to use Equations S1-34 to develop an expression for 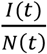 in terms of the parameters *K*_*T*_(*t*) and *P*_*c*_(*t*); and then, with the help of Equation S1-31, express the solution solely in terms of these parameters. We first note that every person within both *N*(*t*) and *I*(*t*) contacts *P*_*c*_(*t*) people in each period Δ*t*. Therefore, in accordance with Equation S1-9, beginning at *t* = 0, *K*_*T*_(Δ*t*)*I*(0)Δ*t* people within the *I*(0)*P*_*c*_(0) group become infected at *t* = Δ*t*; and, concurrently, the number of non-infected people within the *I*(0)*P*_*c*_(0) group becomes *I*(0)*P*_*c*_(0) − *K*_*T*_(Δ*t*)*I*(0)Δ*t* + *I*(0)Δ*P*_*S*_(0). We add the term *I*(0)Δ*P*_*S*_(0) to account for any susceptible people that join or leave the *P*_*c*_(*t*) group; and we assume people who have joined will only be infectable in the next time step.

Apart from the allowance that the number of susceptible people within *P*_*c*_(0) can change, this approach is the same as Kermack and McKendrick’s concept that the susceptible density surrounding an infected person diminishes with each infection transmission. The subtle difference between our conceptualization and Kermack and McKendrick’s is that by using *P*_*c*_(*t*) we mathematically force the growth of the epidemic to be a local phenomenon which can only reduce the susceptible density in direct contact with the infected individual(s). This is consistent with Kermack and McKendrick’s paradigm and has the advantage of eliminating the complexity that they introduced with the implicit assumption that the contacts between all the people declined in proportion to the shrinking number of globally available susceptible people.

In harmony with another of Kermack and McKendrick’s assumptions and the concept of latency in all infectious persons, by specifying the term *K*_*T*_(Δ*t*) as the transmission rate causing the new infections in this first step, we ensure that a newly infected person cannot infect others immediately after they become infected. Therefore, in concert with Kermack and McKendrick’s interpretation that *φ*(0,0) = 0, we assume that *K*_*T*_(*t*) = 0 for *t* ≤ 0.

Since 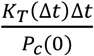 is the fraction of *P*_*c*_(0) the contacting infected people can no longer infect, and 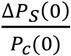 is the fraction of <u>new</u> contacts within *N*(*t*) that can be infected, the factor, 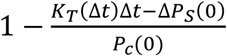, is the fraction of *P*_*c*_ (*t*) that remains susceptible. After a time Δ*t*, therefore,

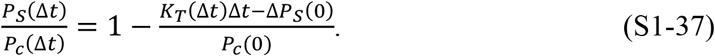

During the next time step, another *K*_*T*_(Δ*t*)*I*(Δ*t*)Δ*t* people within the group *I*(Δ*t*)*P*_*c*_(Δ*t*) become infected and the fraction of susceptible people within 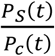 changes by an additional factor,

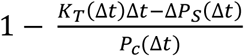. This is expressed mathematically as,

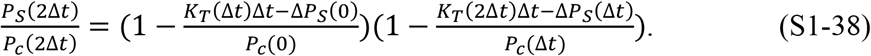

The process repeats itself in each period Δ*t*; and we can write,

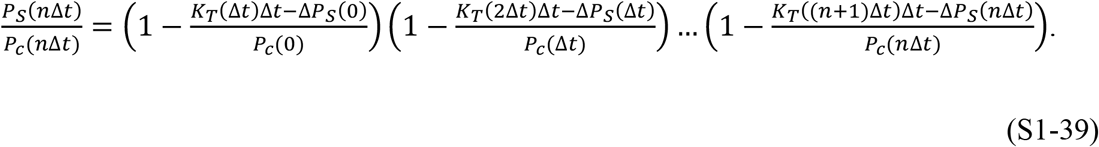

Applying the second thought experiment, we now make the simplifying assumption that a change, Δ*P*_*S*_(*t*), to *P*_*S*_(*t*) will occur much more slowly than the creation of new infections, meaning that in Equation S1-39, Δ*P*_*S*_(*n*Δ*t*) ≪ *K*_*T*_((*n* + 1)Δ*t*)Δ*t*. Therefore, in the remaining analysis we will neglect the Δ*P*_*S*_(*n*Δ*t*) terms. (A situation where ΔP_S_(nΔt) is plausibly growing as fast as K_*T*_(nΔt)Δt indicates an outbreak from the initial epidemic and is explored further in Supplement 3.1.)

Neglecting the Δ*P*_*S*_(*n*Δ*t*) terms in Equation S1-39, using Equation S1-33; and, since by definition, *n*Δ*t* = *t*, as *n* → ∞, Δ*t* → 0, Equation S1-39 becomes,

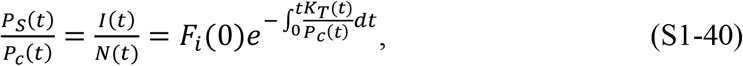

where 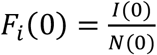; the fraction of *N*(*t*) infected at *t* = 0. We can also relabel *P*_*S*_ (*t*) as *R* _*eff*_ (*t*) because *P*_*S*_(*t*) is the number of additional people each person in *I*(*t*) will eventually infect. Equation S1-40 is the expression that enables us to bring the population interactions into the solution.

### Step 4: The KMES

When Equation S1-40 is substituted into Equation S1-31, N(t) can be expressed in terms of *K*_*T*_(*t*) and *P*_*c*_(*t*),

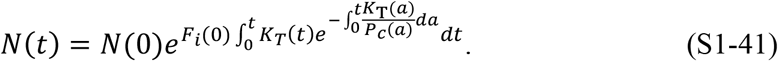

*I*(*t*) is then found by multiplying Equations S1-40 and S1-41 together,

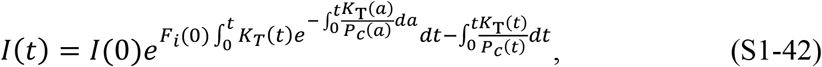

and from the definition of *N*(*t*), *R*(*t*) is,

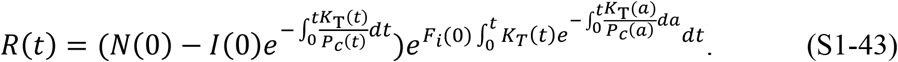

Substituting Equations S1-41 to S1-43 into Equations S-9 through S-11 yields the expression,

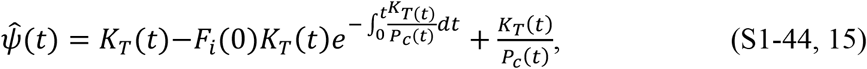

which defines the relationship between 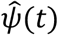, *K*_*T*_(*t*), and *P*_*c*_(*t*).

As a check, this expression for 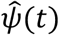 can be substituted into Equations S1-13 and S1-14 to demonstrate their equivalency to Equations S1-41 and S1-42. We therefore conclude that Equations S1-41, -42, and -43 are solutions to Kermack and McKendrick’s integro-differential equations in terms of the transmission and population interaction parameters, *K*_*T*_(*t*) and *P*_*c*_(*t*).

Written out, the complete solution to Kermack and McKendrick’s integro-differential equations (the KMES), in terms of *K*_*T*_(*t*) and *P*_*c*_(*t*) is,

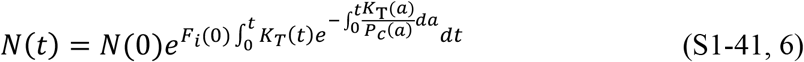

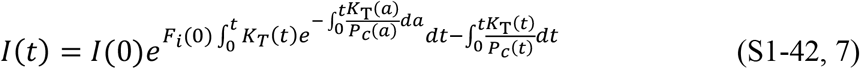

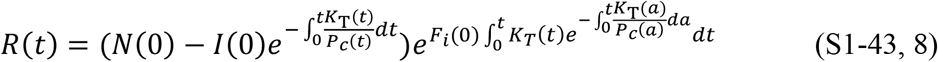

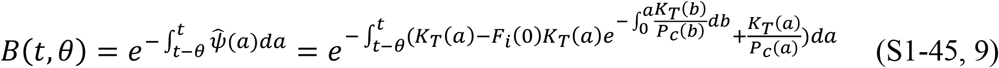

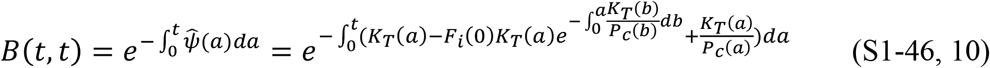

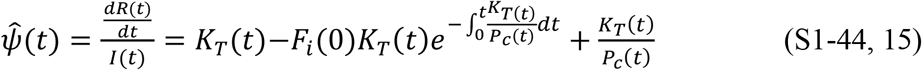

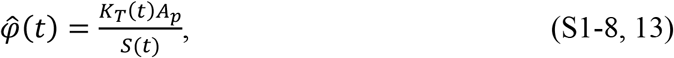

and since *S*(*t*) = *N*_*P*_ − *N*(*t*), rather than portray the progress of the epidemic in terms of susceptibles, *S*(*t*), we henceforth express the KMES in terms of the total cases, *N*(*t*), as in Equation S1-41.

## Supplement 2. Controlling epidemics early

The quantitative mathematical relationships derived from the KMES presented in Section 1 characterize the dynamics of an epidemic and illustrate that strong and early intervention is critical. Equation 37 quantifies that the ultimate number of individuals infected in an epidemic, *N*(∞), will be exponentially dependent on the number of people with whom each person interacts.

The real-world country data provide vivid examples of the consequences projected by the KMES. Both South Korea and New Zealand enacted strong and early interventions compared to other countries (Campbell, C 2020; Field, A 2020), as reflected by their 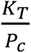 values (Table 2). These strong interventions led to earlier peaks in new cases and to far fewer total cases than in other countries (Figures 5 and 6) in the first few months of the pandemic: the peak number of new cases in both South Korea and New Zealand was 90–99% lower than in other countries, a compelling validation of the explicit statement in the KMES that strong intervention leads to *exponentially* more favorable outcomes.

In the USA, interventions initiated on March 16 began to have an effect around March 23, 2020 (Figure 5B), and the number of new cases on March 23, 2020 (Roser et al 2021) was 46,136. Using the values of ln (*F*_*i*_ (0)*K*_*T*_ (t)) and 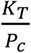 calculated from the data, Equation 37 predicts that had the value of these parameters remained constant, the ultimate number of cases would have been approximately 1.22 million. If the same intervention had been implemented and sustained starting on March 10, when there were 59 times fewer (782) cases (Roser et al 2021), the model predicts that the ultimate number of cases would also have been 59 times lower, or 20,725. Thus, earlier action could have reduced the ultimate number of projected cases by more than 98%. Of course, the projected estimate of approximately 1.22 million total USA cases would only have occurred if the effectiveness of the interventions that were launched on March 16 had been sustained. Unfortunately, a marked reduction in effective interventions occurred in many parts of the USA in mid-April, well before the official reopening of the economy (Elassar 2020). This caused a second surge in new cases in late April and is why the observed data and the model prediction diverge in Figures 5B and 6B.

As shown in Section 1, the KMES via Equation 35 provides an estimate of the time to the peak of new cases, *t*_peak_. Using Equation 35, the values of ln(*F*_*i*_ (0)*K*_*T*_ (t)), and 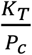 calculated from the data, the predicted peak in new cases in the USA would have occurred near March 24 if the intervention had begun on March 10. Instead, a 6-day delay in effective intervention shifted the initial peak to April 11, 16 days later, as projected, and that peak was much higher (Figure 6B).

As shown, too, in Section 3, epidemic acceleration, the instantaneous potential to change the pace of the epidemic, can be determined at any point in the epidemic and depends on the social containment actions in effect at that time. What is perhaps less apparent, but predicted by the KMES, is that two countries with identical numbers of cases on a given day can, in fact, have different accelerations on the same day, and will, therefore, exhibit different dynamics immediately after that day.

South Korea and New Zealand (Figure 5A and F) had nearly identical case counts when each imposed strong containment measures (204 cases in South Korea on February 21, and 205 in New Zealand on March 25). Their data suggest that their interventions were similarly effective (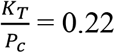 in South Korea and 0.12 in New Zealand; see Table 2). However, since South Korea has a much higher population density than New Zealand ((Worldometers 2021), data in Table 1), it had a much higher number of interactions when the interventions were imposed and, therefore, a higher rate of acceleration, as evidenced by its higher RCO at the time of intervention. Indeed, the rate of change of new cases *was* higher in South Korea than in New Zealand, and the later number of cases in South Korea *was* higher than in New Zealand (Figure 5A and F).

Equation 33 clearly illustrates these lessons. As social distancing is strengthened (lower *P*_*c*_), *R*_*eff*_(*t*) decreases, and the epidemic slows. Early and strong interventions, especially in countries with indigenously high levels of social interaction, are necessary to stop an epidemic in the initial stages. Reopening, enacted too early, can reignite the epidemic, dramatically increasing the number of cases. The astonishing magnitude of the effects, driven by only a few days of delay, derives from the doubly exponential nature of the underlying relationships.

## Supplement 3. Ending an Ongoing Epidemic

We can use the KMES to design measures to end an epidemic in an advanced stage. The management plan is built by first using Equation 62 to predict the number of days a given level of intervention, 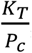, is needed to reduce the new daily cases by a target fraction, *D*_*tf*_.

For example, using Equation 62, we see that a country targeting a 90% reduction of new cases per day (e.g., from 50,000 to 5,000 cases per day, *D*_*tf*_ = 0.1), can attain its target in about 12 days by imposing a containment level of 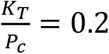. The South Korea and New Zealand data demonstrate that Equation 62 is valid and that 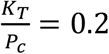 is achievable for this duration. Both countries achieved a value of 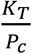 close to 0.2 for the time necessary to produce a 90% reduction. It took 13 days for South Korea (March 3–16) and 15 days for New Zealand (April 2–15) to reduce their new cases by 90% between the dates shown.

Returning to the planning example, after achieving the initial 90% reduction, a reasonable next step might be to relax social containment to a level that allows the economy to remain viable, while preventing the epidemic from erupting again. We can again find the level of 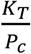 necessary to achieve a chosen target using Equation 62. If an additional 90% reduction in new cases per day is desired, and a period of 90 days is tolerable for that reduction, then a new level of approximately 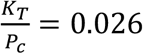 is needed. This equates to a 90-day period during which each person can be in contact with ten specific people, in an infectable way. Note that this is three times *less* stringent than the original USA shutdown level in April 2020 as shown by the level of 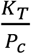 calculated for the United States in that period (Table 2). Thus, with a well-planned approach, a country can reduce its new daily cases by 99% in approximately 100 days, enabling the country to control, and essentially end the epidemic, while simultaneously maintaining economic viability.

If even 0.026 is too restrictive, we can choose a still lower 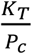, but it must be large enough to avoid a new outbreak. A lower bound for the new value of 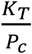, high enough to prevent an outbreak, can be found using Equation 59.

We can easily monitor the progress of interventions using the RCO, as the curve for South Korea illustrates (Figure 5A). Had this country maintained the initial level of distancing measures, the data would have followed the initial slope. However, the actual data departed from the slope, heralding failures in (or relaxation of) social distancing, which were later documented to have occurred during the indicated time frame (Campbell 2020) (circled data, Figure 5A). Because it summarizes epidemic dynamics, we can use the RCO to continuously determine the effectiveness of implemented measures and whether they need adjustment.

### Supplement 3.1 Outbreaks

We can see from Equation 50 that if the social interventions are strengthened (lower *P*_*c*_) the slope of the RCO curve will steepen and if the interventions are relaxed, the slope will become shallower. Therefore, if the value of *K*_*T*_(*t*) does not change due to a change in the disease transmissibility, the RCO is a metric for monitoring the population interactions. It is also clear that, under the assumptions used to develop the KMES, 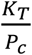 must always be greater than zero, and the RCO slope can never become positive. However, this only remains true if three conditions remain true: 1) immunity persists, 2) no new infections are introduced from outside the area, 3) Δ*P*_*S*_(*n*Δ*t*) is an order of magnitude smaller than the new infections, *K*_*T*_(*n*Δ*t*)Δ*t*. We call the latter two conditions the assumption that the epidemic is contiguous.

If new infections are introduced into a portion of the population that has thus far been disconnected from the previously infected area, and therefore, has only susceptible people, then the assumption of contiguousness does not hold. This is a common situation when infected people travel from an infected area to a previously uninfected area and cause an outbreak.

In this case, we will begin with Equation S1-35 and assume that the entirety of the change in *P*_*c*_(*t*) during the time Δ*t* is with uninfected new contacts. That is, Δ*P*_*c*_(*t*) = Δ*P*_*S*_(*t*); and Equation S1-35 becomes,

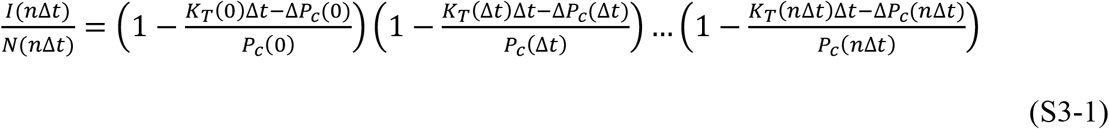

and then, since by definition, *n*Δ*t* = *t*, as *n* → ∞, Δ*t* → 0, Equation S3-4 becomes,

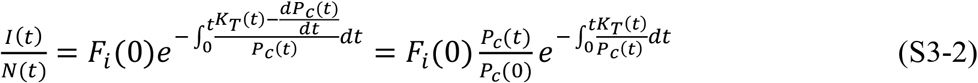

The equations for *N*(*t*), *I*(*t*), and *R*(*t*) are then the following:

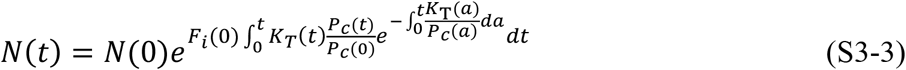

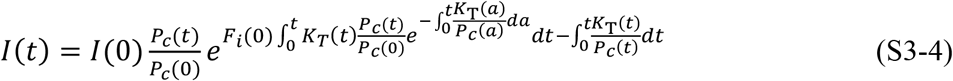

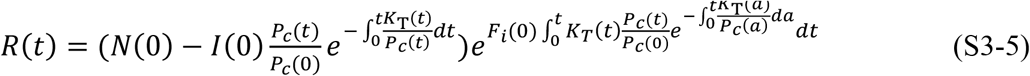

As an alternative, to predict the number of cases in an epidemic affected by an outbreak, we can modify Equation 28. Assuming that *t*_0_ = 0, *N*(0) = 1, and introducing the notation *P*_*cx*_ where *x* denotes the number (order in time) of the outbreak, Equation 28 can be written as:

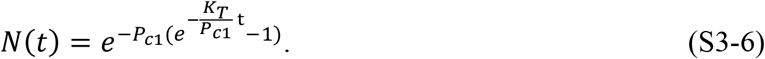

If a new outbreak occurs in a previously unaffected area of a country, then Equation S3-6 can be modified as follows:

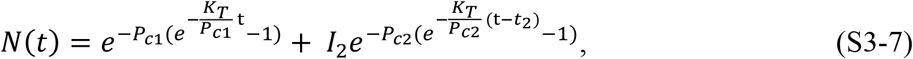

where *I*_2_ is the number of infectious people who initiated the new outbreak, *P*_*c*2_ is the social interaction parameter in the new outbreak area, and *t*_2_ is the time the new outbreak occurs. We have assumed that the disease transmissibility remains the same throughout this illustration. If the transmissibility changes in a subset of the population, then a similar formulation, using the notation, *K*_*Tx*_, can be utilized to track the populations with the new transmissibility.

Equation S3-7 can be written in a general form as

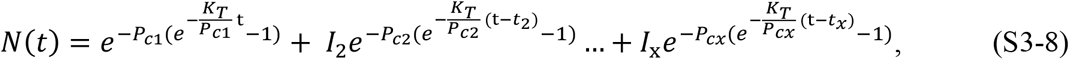

where *x* denotes the outbreak number and t > *t*_2_ > *t*_3_ > ⋯ > *t*_*x*_. For each outbreak *t*_*x*_, *P*_*cx*_, and *I*_*x*_ need to be determined independently.

While an epidemic is underway, we can detect an outbreak by monitoring the slope of the RCO curve. A positive slope detected in an RCO curve indicates that an outbreak has occurred. This is an indication that immediate action, within days, is required from policy makers to strengthen intervention measures and prevent the outbreak from overwhelming prior progress in controlling the epidemic. By monitoring the RCO curve, we can also detect if the disease changes its transmissibility through mutation. In this situation, a proper fit of the parameters in Equation 28 is not possible and a modification of *K*_*T*_ is required to accommodate the change.

## Supplement 4: List of Equations

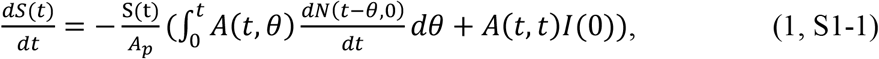

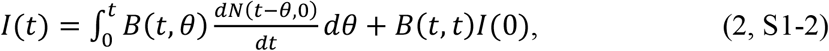

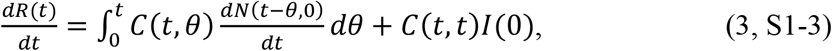

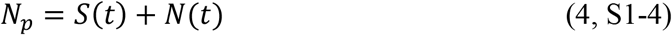

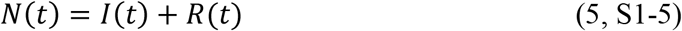

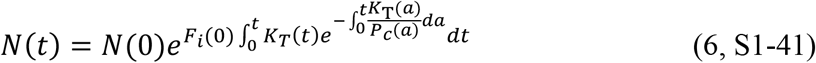

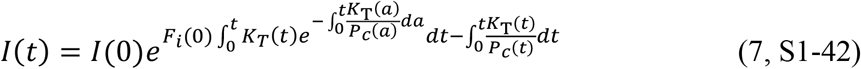

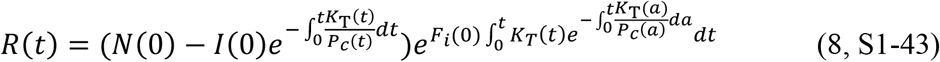

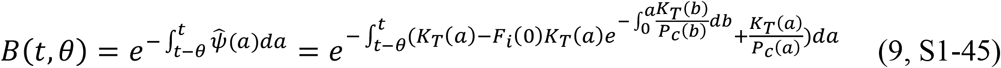

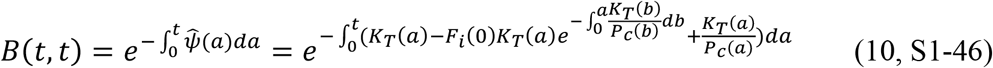

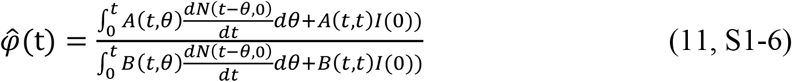

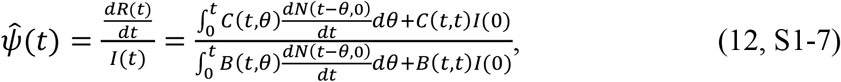

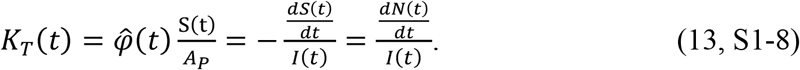

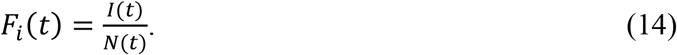

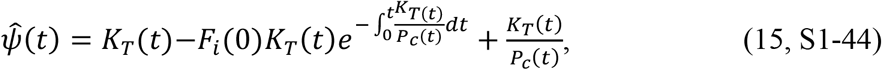

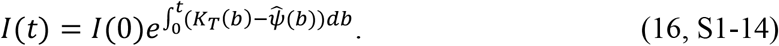

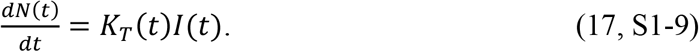

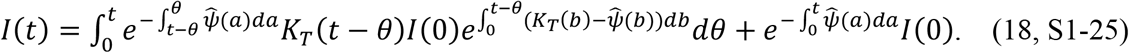

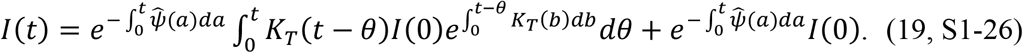

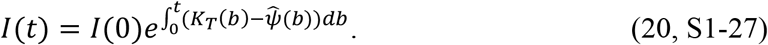

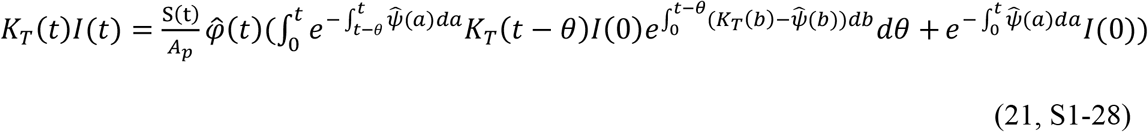

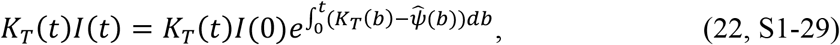

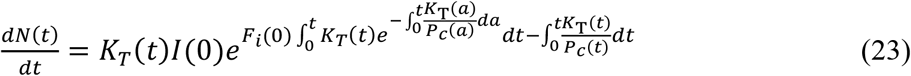

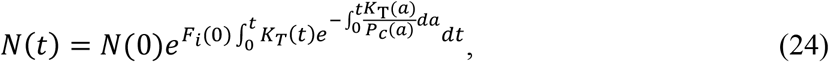

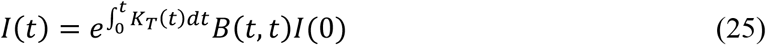

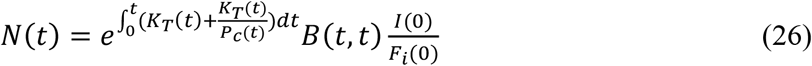

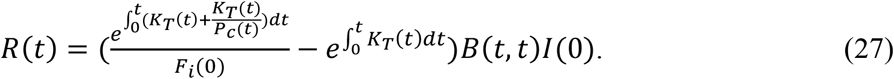

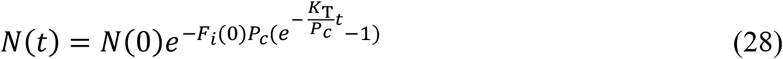

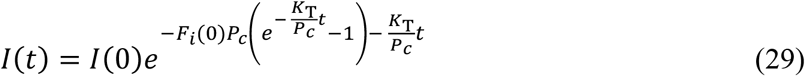

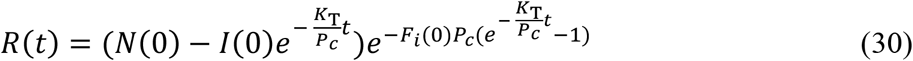

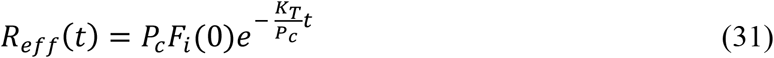

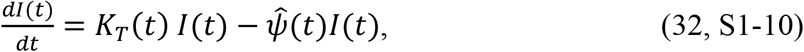

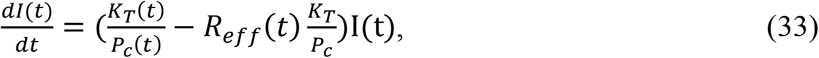

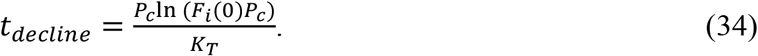

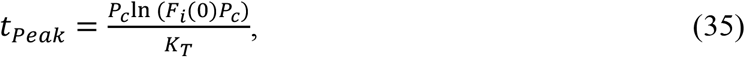

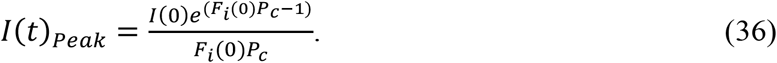

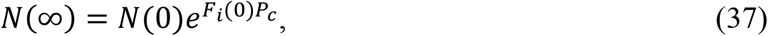

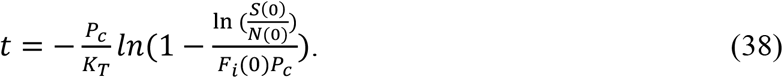

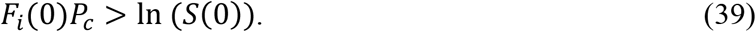

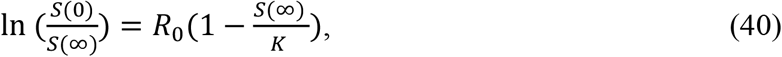

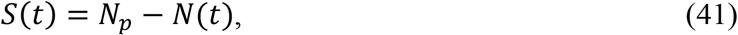

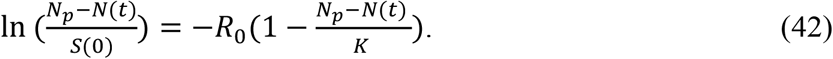

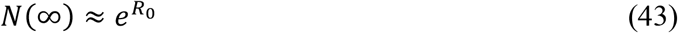

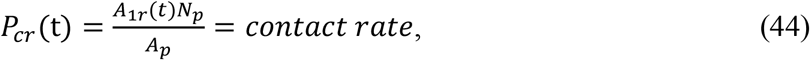

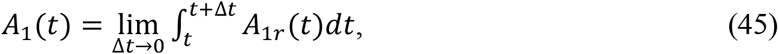

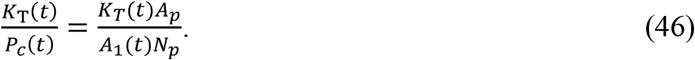

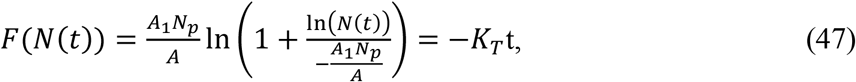

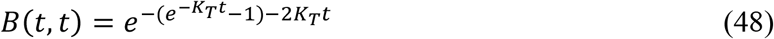

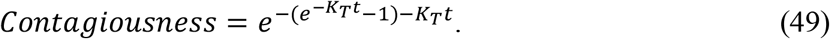

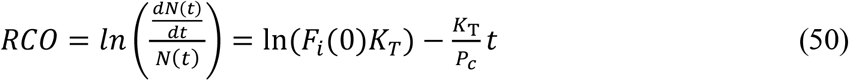

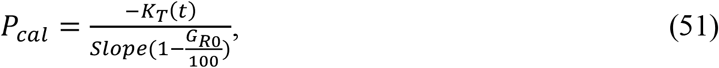

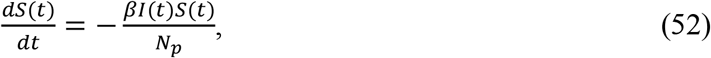

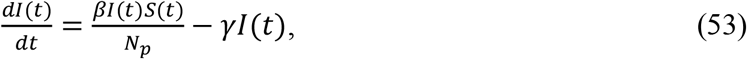

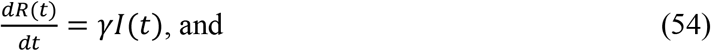

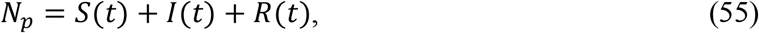

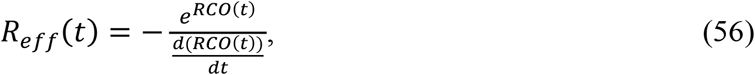

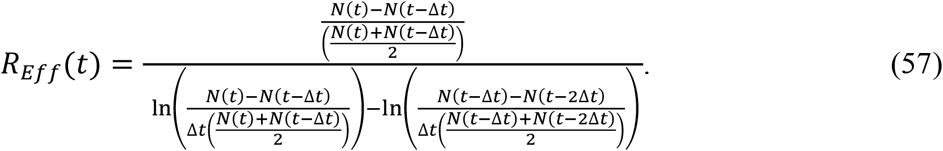

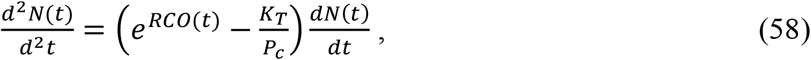

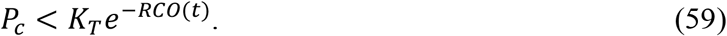

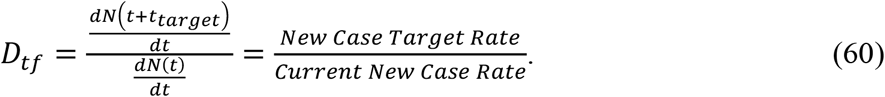

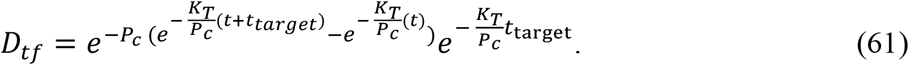

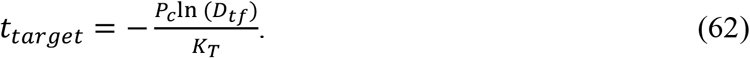

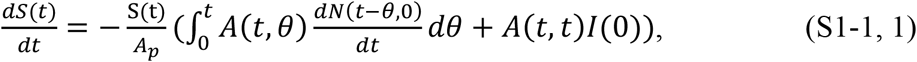

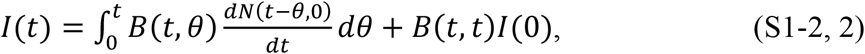

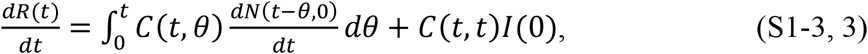

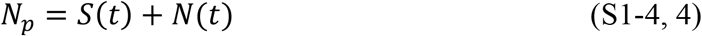

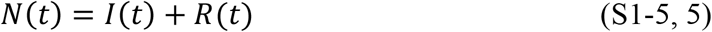

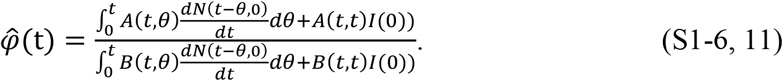

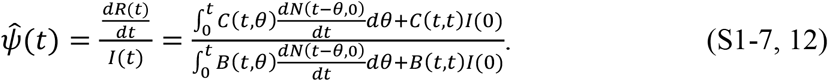

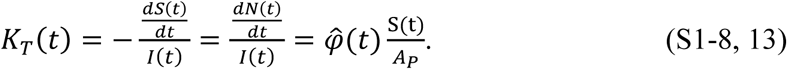

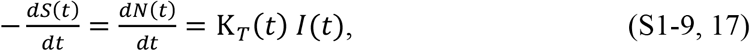

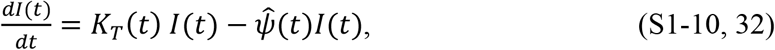

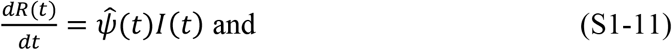

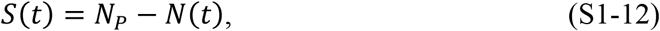

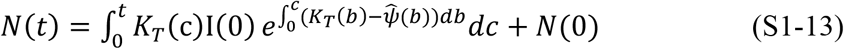

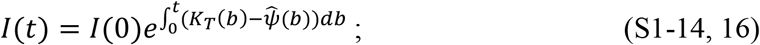

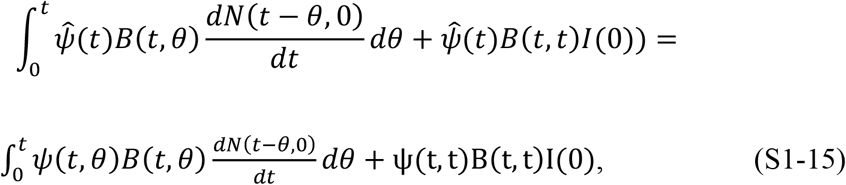

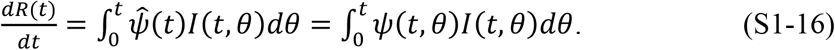

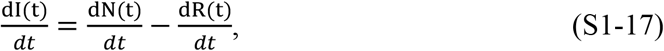

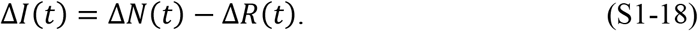

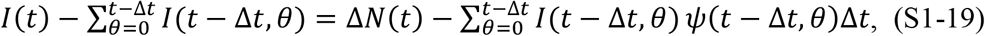

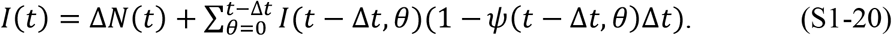

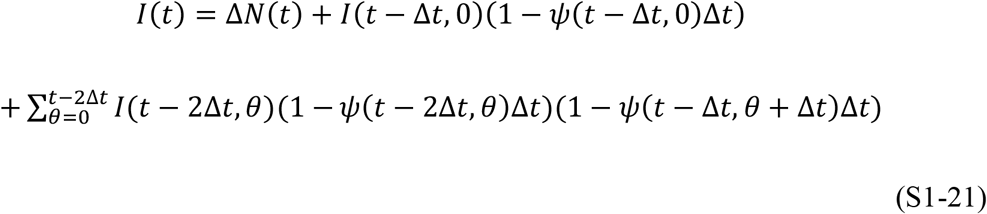

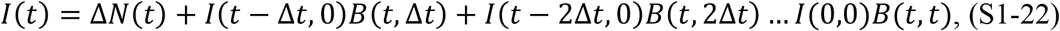

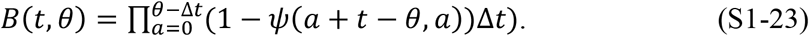

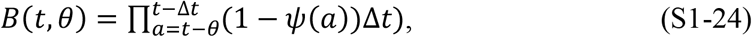

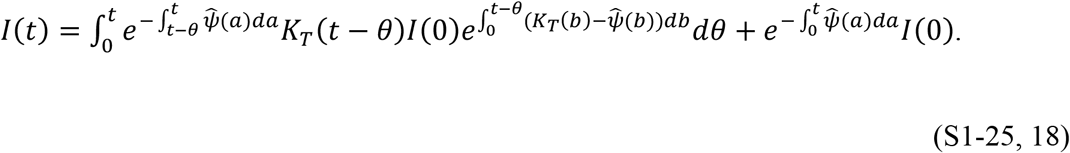

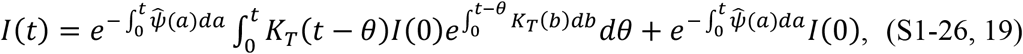

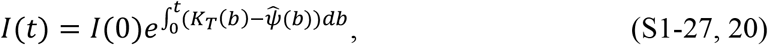

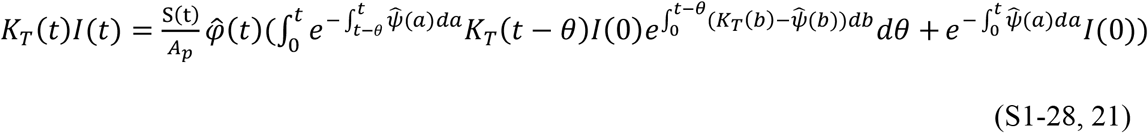

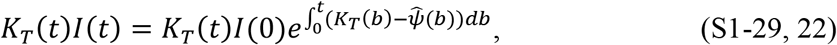

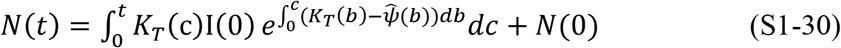

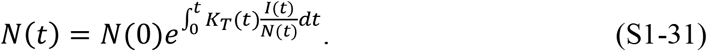

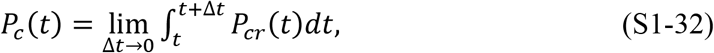

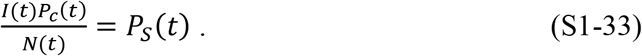

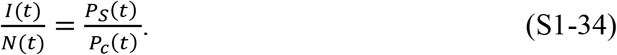

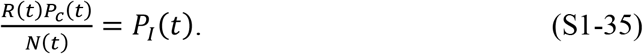

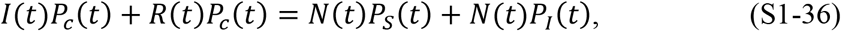

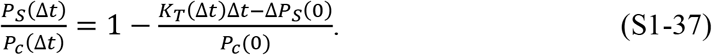

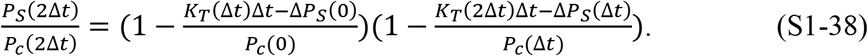

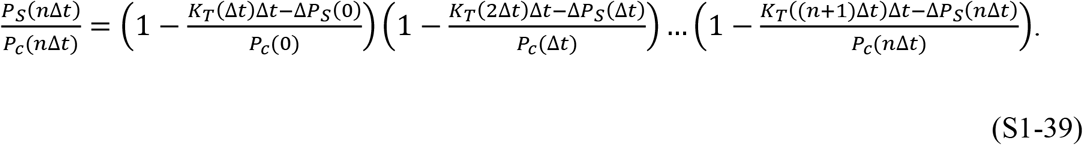

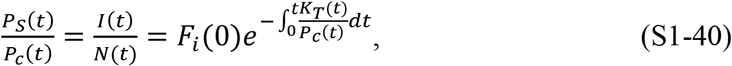

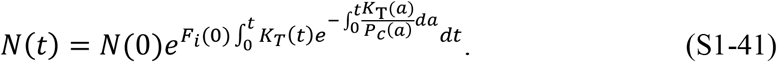

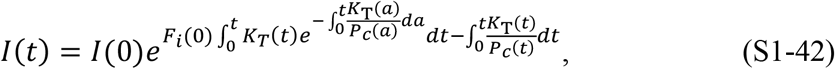

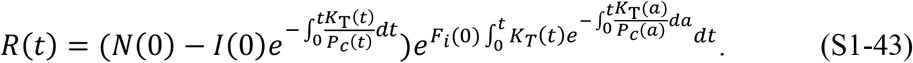

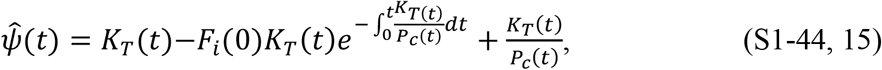

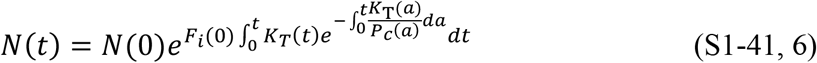

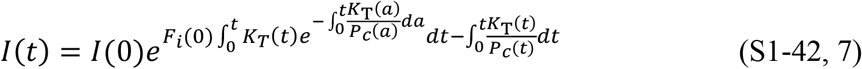

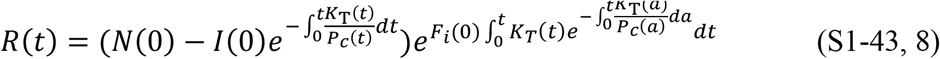

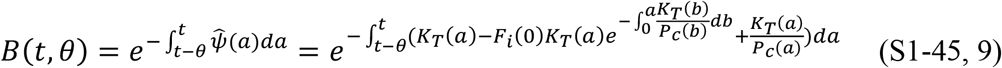

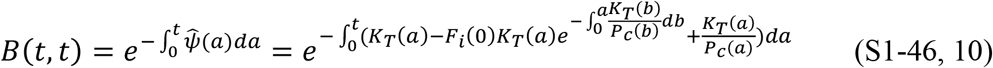

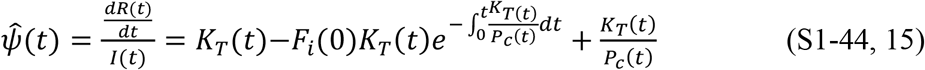

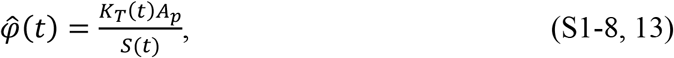

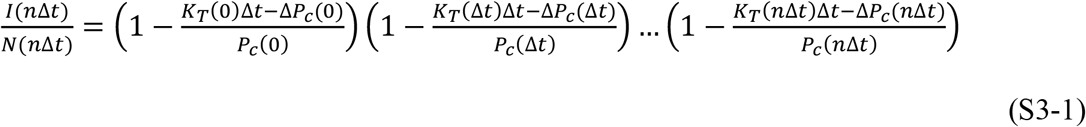

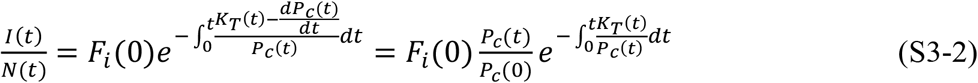

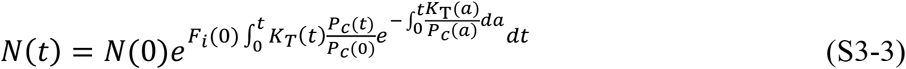

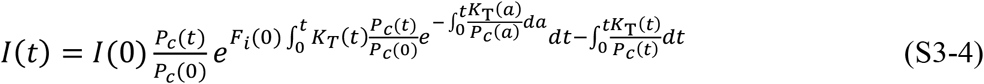

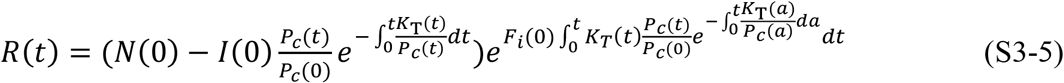

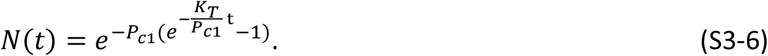

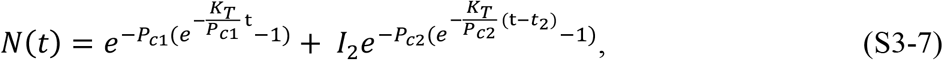

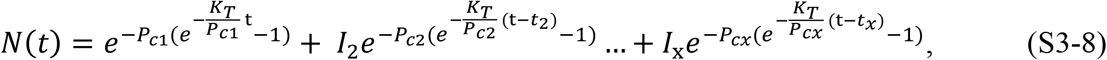

## Notes

### Competing Interest Statement

The authors have declared no competing interest.

### Funding Statement

This study did not receive any funding

